# A systematic review and meta-analysis of non-workplace interventions to reduce time spent sedentary in adults

**DOI:** 10.1101/2021.05.27.21256673

**Authors:** Jessica Faye Hall, Rekesh Corepal, Thomas F Crocker, Natalie Lam, Louisa-Jane Burton, Karen Birch, Gill Carter, David J Clarke, Coralie English, Amanda J Farrin, Claire Fitzsimons, Jennifer Hall, Ivana Holloway, Seline Ozer, Rebecca Lawton, Gillian Mead, Sarah Morton, Anita Patel, Anne Forster

## Abstract

**Background:** Sedentary behaviour has been the focus of considerable clinical, policy and research interest due to its detrimental effects on health and wellbeing. This systematic review aims to (1) develop a more precise description of different categories of interventions that aim to reduce sedentary time in adults by identifying specific components that form an intervention; (2) explore the effect of different categories of interventions in reducing time spent sedentary in adults.

**Methods:** Ten electronic databases, websites of relevant organisations (e.g. the Sedentary Behaviour Research Network), and relevant reviews were searched. *Inclusion criteria:* Randomised controlled trials (RCTs), including cluster and randomised cross-over trials, in the adult population (clinical and non-clinical). Any study including a measure of sedentary behaviour was included even if reducing sedentary behaviour was not the primary aim. *Exclusion criteria:* Interventions delivered in schools, colleges, or workplaces; studies investigating the immediate effects of breaking up sitting time as part of a supervised (usually laboratory-based) intervention. Two review authors conducted data extraction and quality assessment (GRADE approach).

**Results:** Searches identified 39,223 records, of which 85 studies met the inclusion criteria and were included in the review. Interventions shown to significantly reduce time spent sedentary were those which incorporated the provision of information, education, or support (advice/recommendations), in conjunction with either counselling (mean difference: -52.24 minutes/day; 95% CI: -85.37 to -19.10) or a form of structured/prescribed physical activity (standardised mean difference: -0.15; 95% CI: -0.23 to -0.07). However, this positive effect was not maintained at follow-up. No interventions were shown to break up prolonged sitting.

**Conclusions:** This review presents a novel way of categorising interventions according to the types of components they comprised. There is evidence that interventions might be effective in reducing time spent sedentary immediately post-intervention. There were limited studies measuring sustained behaviour change.

## Background

Sedentary behaviour, defined as any waking behaviour characterised by low energy expenditure ≤1.5 Metabolic Equivalents (METs) while in a sitting, lying or reclining posture (1), is the focus of considerable clinical, policy and research interest due to its detrimental effects on health, wellbeing and economic burden (2-5). There is evidence for the negative impacts of sedentary behaviour on many health parameters (4, 6, 7), including reduced physical function (8, 9), increased symptoms of depression (10) and anxiety (11), and increased risk of cardiovascular disease (12, 13) and premature mortality (14-17).

Recent estimates suggest that adults spend a large proportion of their day in sedentary behaviours. One review found that the median self-report of daily sitting time was 5.5 hours per day (range from 3.8 hours per day to 7.6 hours per day), and was even higher (median 8.2 hours per day; range from 4.9 to 11.9 hours per day) when looking at studies that measured with devices (18).

Earlier research reported that the negative effects of sedentary behaviour are independent of the level of daily moderate to vigorous physical activity (MVPA) undertaken (1, 19, 20). However, a meta-analysis conducted by Ekelund et al. (21) suggested very high levels of MVPA (approximately 60 to 75 minutes per day) may eliminate the increased risk of early death associated with long periods of sitting time (>8 hours per day). Globally, however, only 27.5% people undertake the current recommended levels of physical activity (>150 minutes per week of moderate physical activity or >75 minutes vigorous physical activity per week, or an equivalent combination of the two) (22). Therefore, it might be more feasible for the majority of the population to reduce sedentary behaviour than to increase physical activity to levels which might attenuate the effects of sedentary behaviour.

Previous reviews have identified some types of interventions which have successfully reduced sedentary behaviour (23-27). In the systematic review and meta-analysis by Prince et al. (23), interventions focusing on sedentary behaviour were found to be more effective at reducing sedentary behaviour than interventions aimed at reducing sedentary behaviour in conjunction with increasing physical activity, or interventions focused on increasing physical activity only. In Shrestha et al’s. (28) systematic review and meta-analysis, interventions targeting specific sedentary behaviours, such as leisure sitting time and TV viewing, were shown to be effective. However, Shrestha and colleagues could not conclude which type of interventions among educational, multi-component, TV-restricting, and workplace were the most effective due to the heterogeneity of interventions tested and low number of studies of each intervention type.

Martin et al. (24) recommended more precise descriptions of interventions in future systematic reviews, to identify potential single or multiple component interventions to reduce sedentary behaviour. Subsequent reviews have gone some way towards categorising interventions; however have either tended to use quite broad categories (e.g. adult or children interventions, lifestyle interventions, and behavioural or environmental interventions) (28-30) or have focussed only on behaviour change techniques and functions used (25). No systematic reviews have yet performed meta-analyses of studies according to their key intervention components (including the specific combinations of multiple components), to identify which are more effective for reducing sedentary behaviour in adults.

To address this gap, this systematic review and meta-analysis provides an updated and rigorous review of randomised controlled trials in adults, with the aim to:

- Develop a more precise description of different categories of interventions to reduce time spent sedentary by identifying specific components (and combinations of components) that form an intervention;
- Explore the effect of these different categories of interventions in reducing time spent sedentary (which includes number of breaks in sedentary behaviour) when compared to no treatment, usual care, attention controls, and alternative treatments.

This review is designed to offer guidance for use in the development of future interventions aimed at reducing sedentary behaviour by highlighting the intervention components, and possible combinations of components that are effective in reducing sedentary behaviour in adults. In particular, the review will contribute to a National Institute for Health Research programme grant for the development and evaluation of strategies to reduce sedentary behaviour in patients after stroke. Taking into account the fact that the majority of strokes occur in adults aged 65 years and over (31), interventions that take place in schools, colleges, universities and workplaces were excluded from the review as they are less applicable to our population of interest.

## Methods

### Protocol and registration

Reporting of the systematic review is guided by Preferred Reporting Items for Systematic Review and Meta-analysis (PRISMA) guidance (32), as shown in the PRISMA checklist (Additional file 1). To provide full transparency and avoid study duplication, the systematic review was prospectively registered with PROSPERO (Prospective Register of Systematic Reviews); registration number: CRD42018083751.

### Study selection criteria

#### Study design

Randomised trials, including cluster and randomised cross-over trials. Studies where the interventions were compared to a control group (including no active treatment, wait list control, attention control, usual care) or alternative treatments were included. Studies were excluded where the main aim was to investigate the immediate effects of breaking up sitting time as part of a supervised (usually laboratory based) intervention.

#### Population

Studies involving adults only (as defined in the study), including both clinical and non-clinical populations.

#### Interventions

Any interventions for which a measure of sedentary behaviour was an outcome of interest including, for example, interventions which primarily aim to increase physical activity or promote weight loss but which also lead to changes in sedentary behaviour. Studies which evaluated interventions delivered primarily in schools, colleges, universities and workplaces were excluded.

#### Outcomes

To be included in the analysis, studies must include an outcome measuring time spent sedentary or the number of breaks in sedentary behaviour. The outcome could be measured using devices (e.g. accelerometer, inclinometer) or self-report questionnaires/scales (e.g. International Physical Activity Questionnaire (IPAQ) (33), Marshall Sitting Questionnaire (34)).

### Data sources and searches

#### Data sources

In collaboration with information specialist colleagues, comprehensive search strategies were developed. Publication status, date and/or language restrictions were not applied. In October 2017, ten electronic databases (MEDLINE, Medline in Process & Other Non-Indexed Citations, Embase, Cochrane Database of Systematic Reviews, CINAHL (Cumulative Index to Nursing and Allied Health Literature), PsycINFO, SportDiscus, Cochrane Central Register of Controlled Trials, Web of Science, ClinicalTrials.gov), The Sedentary Behaviour Research Network, the World Health Organisation and US Centres for Disease Control and Prevention websites, were searched. To identify further published, unpublished, and ongoing trials, we examined reference lists of previous reviews.

#### Search strategy

The search strategy for MEDLINE is shown as an exemplar in Additional file 2. This search strategy was adapted for use with the other electronic databases. Endnote x7 reference management software was used to manage and record data during the screening process.

#### Selection process

Two review authors (JFH, RC) independently screened all identified titles and abstracts and retained any that were potentially relevant. Full-text articles of these were obtained and assessed against the review’s inclusion criteria independently by the same two authors. Papers that did not report any measures of time spent sedentary or time spent sitting were excluded. Disagreements were resolved through the involvement of a third reviewer (AF).

#### Data extraction and quality assessment

Two review authors (JFH, RC) piloted the data extraction form with an included study independently and discussed any discrepancies with extracted items. Following this, RC and JFH independently double-extracted 50% of the studies, then RC extracted the remaining studies. A full list of extracted items can be obtained from PROSPERO (CRD42018083751).

Two reviewers (NL, EG) independently assessed each trial’s risk of bias in six domains (random sequence generation, allocation concealment, blinding of participants and personnel, blinding of outcome assessment, selective reporting, and incomplete outcome data) using the Cochrane Collaboration’s tool for assessing risk of bias (35). For each domain, a judgement of high, low or unclear bias was made. Any discrepancies were resolved with involvement from a third reviewer (RC).

The quality of evidence for each study was independently assessed by two reviewers (NL assessed 100% of the studies, RC double assessed 50% of the studies, JFH double assessed the remaining 50% of studies). Quality of evidence for the outcomes of ‘total time spent sedentary’ and ‘number of breaks in sedentary behaviour’ were assessed using the GRADEpro software (36), following the Grading of Recommendations Assessment, Development, and Evaluation (GRADE) system (37). Overall quality scores were determined using the following criteria: risk of bias, inconsistency, indirectness, imprecision, and publication bias. For each outcome, a judgement of high, moderate, low, or very low quality was applied.

### Data synthesis and analysis

#### Categorising interventions before conducting the meta-analysis

The interventions described in the included studies were diverse; therefore, categories were developed to group the interventions before undertaking the meta-analysis. The categories described the key components of the intervention, rather than categorising in terms of what the intervention intended to achieve e.g. a reduction in sedentary behaviour, or the methods used e.g. ‘group-based’. To increase the rigour of categorisation, a group of four researchers (JFH, RC, EG, AE) engaged in a three-stage process.

##### Stage One

Each researcher individually allocated all interventions to initial categories that were developed independently.

##### Stage Two

Consensus-based discussions between all the researchers were used to refine the categories.

##### Stage Three

The final categories were agreed by consensus between researchers and category descriptions were produced.

##### Stage Four

All interventions were allocated to one of the categories finalised in Stage Three through consensus-based discussions between the researchers.

#### Synthesis of results

Study characteristics, participant characteristics and intervention results were extracted and summarised in tabular form. Meta-analyses were conducted only where trials compared the differences between the intervention and control groups for the following outcomes: time spent sedentary (minutes per day) and breaks in sedentary behaviour (number of events per day). Where time spent sedentary was presented as hours per day, it was converted to minutes per day. Meta-analyses were performed using the generic inverse variance method and a random effects model in Review Manager Version 5.3.

Interventions were categorised using the consensus exercise described above, to compare their effectiveness for each outcome. For studies with multiple intervention groups within the same category, the intervention arms were pooled into one group to create a single pair-wise comparison, as recommended (38). In this review, we extracted data for measurements taken immediately post-intervention (“intervention end”), and the last follow-up where there was further follow-up beyond this time (“final follow-up”). Studies using devices to measure sedentary behaviour were analysed separately from studies using self-report measures. Effect sizes were estimated using mean differences (MD) for devices, or standardised mean differences (SMD) for self-report data (35).

Heterogeneity was measured using the I^2^ statistic, with the aim to explore heterogeneity with subgroup analyses when any meta-analysis detected I^2^ ≥50%. When relevant data were not reported, study authors were contacted by email to request the required information. In cross-over trials, only data from the first phase of the trial was used in meta-analyses to guard against carry-over effects. Where cluster RCTs presented an estimate of the effect that accounts for the cluster design, this was used. Where no estimate of the effect was available, an intra-cluster correlation coefficient (ICC) was used (35). Where the ICC for an outcome was not available, an ICC of 0.01 was used (24).

## Results

### Study selection

The PRISMA flow diagram is shown in Figure 1. The search strategy identified 39,223 articles through database searches, and 1,010 articles were added after screening reference lists. After duplicates were removed (n = 11,755), 28,478 titles and abstracts were screened, and 27,896 articles were excluded as they did not meet the pre-defined eligibility criteria. Following this, 582 full texts of articles were assessed, of which 499 articles were excluded (Figure 1). In total, 83 full-text articles were included, which comprised 85 studies (two articles contained data from two RCTs each (39, 40)). All studies were reported in English. Sufficient data were available for 55/85 studies to be included in the meta-analyses.

**Figure 1:**
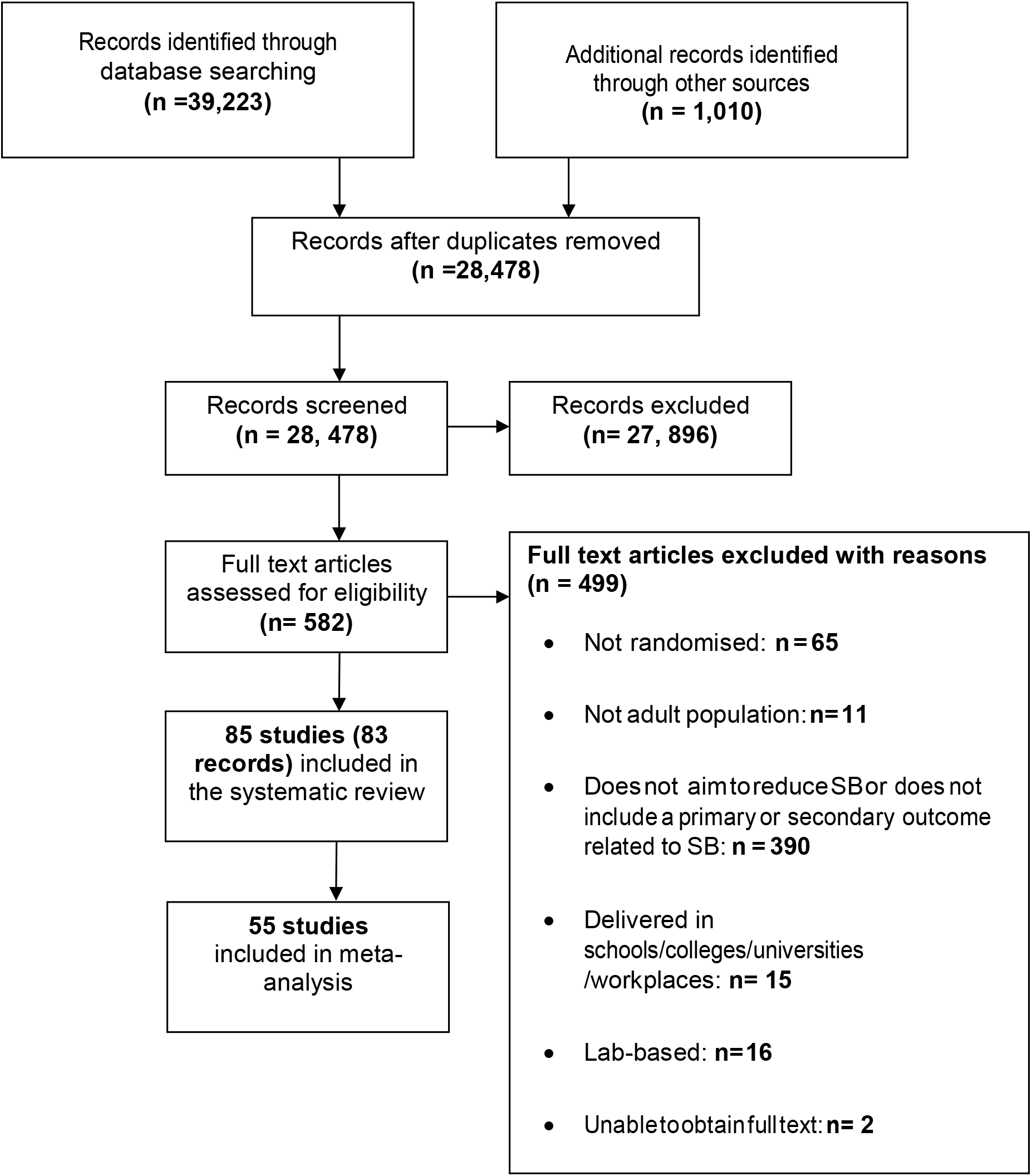
PRISMA flow diagram.

### Study characteristics

The Table of characteristics for included studies is shown in Additional file 3; 85 studies from 83 articles were included. Eighty of the 85 included studies evaluated interventions using RCTs (39-116), and five studies used cluster RCTs; clusters included weight loss groups (117), local government areas (118), senior centres (119), GP practices (120), or intellectual disabilities community-based organisations (121) as the unit of randomisation. Thirty-nine studies were conducted in Europe (41, 43, 44, 47, 48, 51, 52, 54-58, 65-67, 69, 73, 75, 78, 79, 81, 82, 89, 94-97, 99, 100, 103, 105, 110-112, 114, 116, 120-122); 32 were from North America (39, 40, 42, 45, 50, 53, 59, 60, 62, 63, 68, 71, 74, 76, 77, 80, 83-85, 92, 93, 98, 101, 106-109, 115, 117, 119); ten were from Australia (46, 49, 61, 64, 72, 87, 91, 104, 113, 118); and four were from Asia (70, 86, 88, 102). Seven studies used a no intervention control group (48, 49, 64, 65, 81, 85, 108); control participants were asked to maintain their usual lifestyle or were provided usual care in 24 studies (41, 43, 44, 46, 54-58, 66, 69, 70, 75, 90, 91, 93, 96, 99, 101, 102, 110, 111, 113, 118); eleven studies used a wait-list control arm (39, 50, 62, 72, 80, 83, 87, 89, 117, 121); 16 studies used an attention control arm (42, 51, 53, 61, 63, 68, 73, 77, 79, 84, 86, 92, 95, 98, 109, 119); eleven studies used a minimal intervention (47, 60, 76, 78, 88, 97, 103-105, 112, 115) and 16 studies compared more than one intervention with each other without the use of a control (40, 45, 52, 55, 59, 74, 82, 88, 90, 94, 100, 106, 107, 116, 120).

Fourteen studies focused on reducing sedentary behaviour (41, 47, 53, 60-62, 81, 92, 107, 110-112, 117, 119); 34 focused on increasing physical activity (43, 48, 51, 54, 55, 59, 64-66, 68, 71, 73-76, 79, 80, 83, 84, 88-91, 95, 97, 98, 102-105, 108, 114-116); 12 focused on both reducing sedentary behaviour and increasing physical activity (42, 44-46, 56, 57, 67, 77, 86, 94, 113, 121) and 25 focused on other behaviours such as diet, smoking, or alcohol intake in addition to sedentary behaviour and/or physical activity or both (39, 40, 49, 50, 52, 58, 63, 69, 70, 72, 78, 82, 85, 87, 93, 96, 99-101, 106, 109, 118, 120). Sedentary behaviour was measured using devices only (34/85 studies), self-report measures only (39/85 studies), or a combination of both devices and self-report measures (12/85 studies). Regarding the devices, 43/46 studies used accelerometers, and 3/46 used devices attached to the TV to monitor screen time. The most commonly used devices were ActiGraphs™ and ActivPALs™. Self-report measures were more diverse, with the use of various activity logs and questionnaires, e.g. International Physical Activity Questionnaire (IPAQ) (33), Yale Physical Activity Survey (YPAS) (123), past-week Modifiable Activity Questionnaire (PWMAQ) (124), Harokopio Physical Activity Questionnaire (125), Marshall Sitting Questionnaire (34), and Sedentary and Light Intensity Physical Activity Log (126).

In total, 33,074 participants took part in the 85 included studies. The median sample size was 102, with a range from 20 (110) to 12, 287 participants (65). On average, 66% of participants were female. The proportion of females in the studies of mixed sexes ranged between 13% and 92%; five studies included only male participants (39, 73, 87, 95, 114), and 20 studies included only female participants (39, 42, 58, 63, 68, 70, 71, 74, 79, 82, 93, 96, 98, 99, 103, 104, 109, 115, 117, 118). The median of the mean age of study participants was 52 years, ranging from 18 years (73, 95) to 82 years (91).

Twenty-four studies targeted overweight participants (39, 40, 44, 47, 50, 57, 58, 63, 69, 75, 76, 82, 83, 87, 92-94, 96, 101, 107, 117, 120); eleven studies targeted participants with Type 2 diabetes (44, 52, 56, 57, 86, 102), gestational diabetes (70), or those at risk of Type 2 or gestational diabetes (47, 68, 78, 101). Other clinical conditions used as inclusion criteria in studies included rheumatoid arthritis (110, 111), chronic obstructive pulmonary disease (48, 54, 113), multiple sclerosis (84), dementia (119), depression (67), a history of myocardial infarction (90), cancer (72, 98, 104), and stroke (61).

### Intervention categorisation

Fourteen categories of interventions were identified by the categorisation process, including three single-component, and eleven multi-component interventions (see Table 1).

**Table 1:**
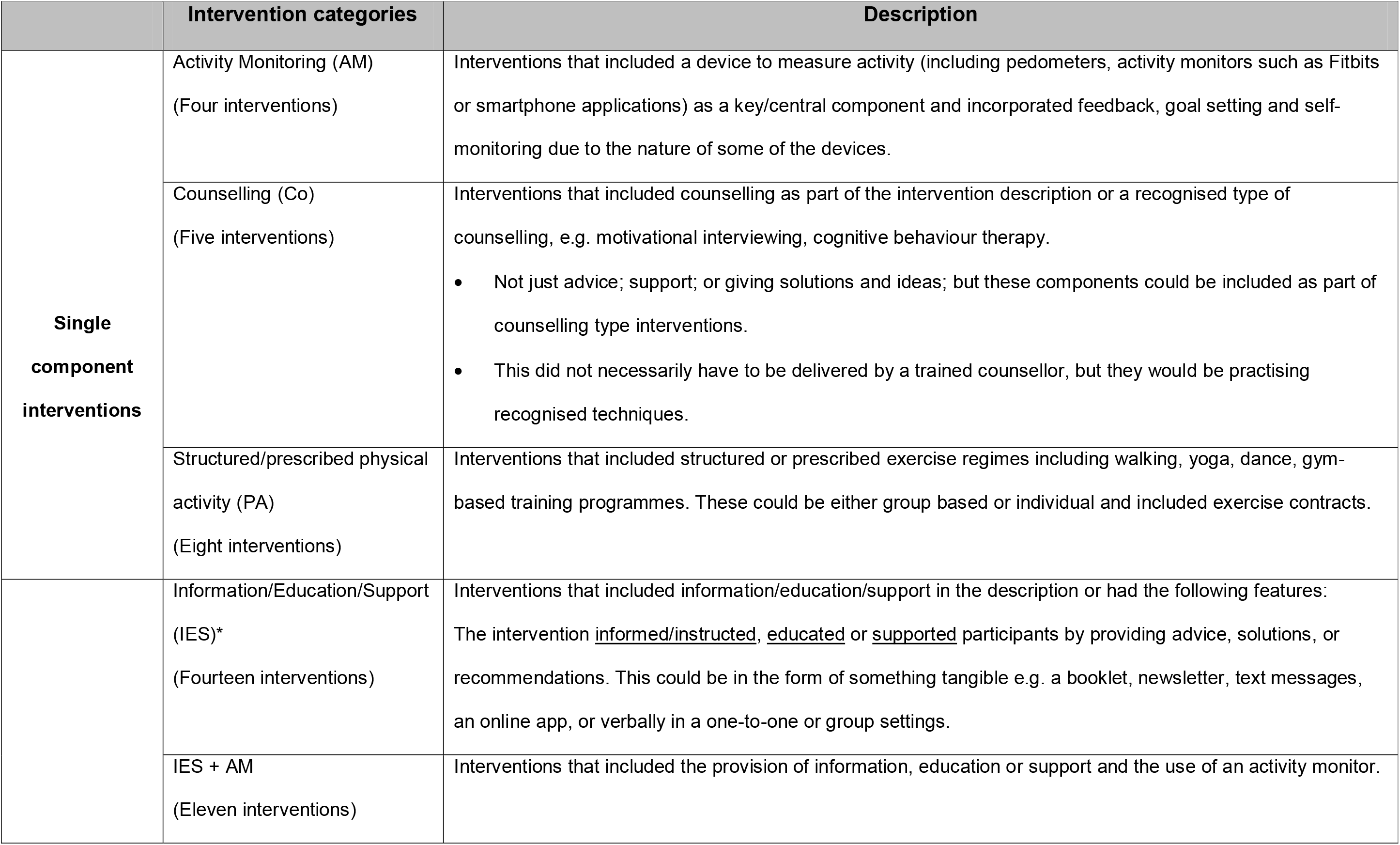
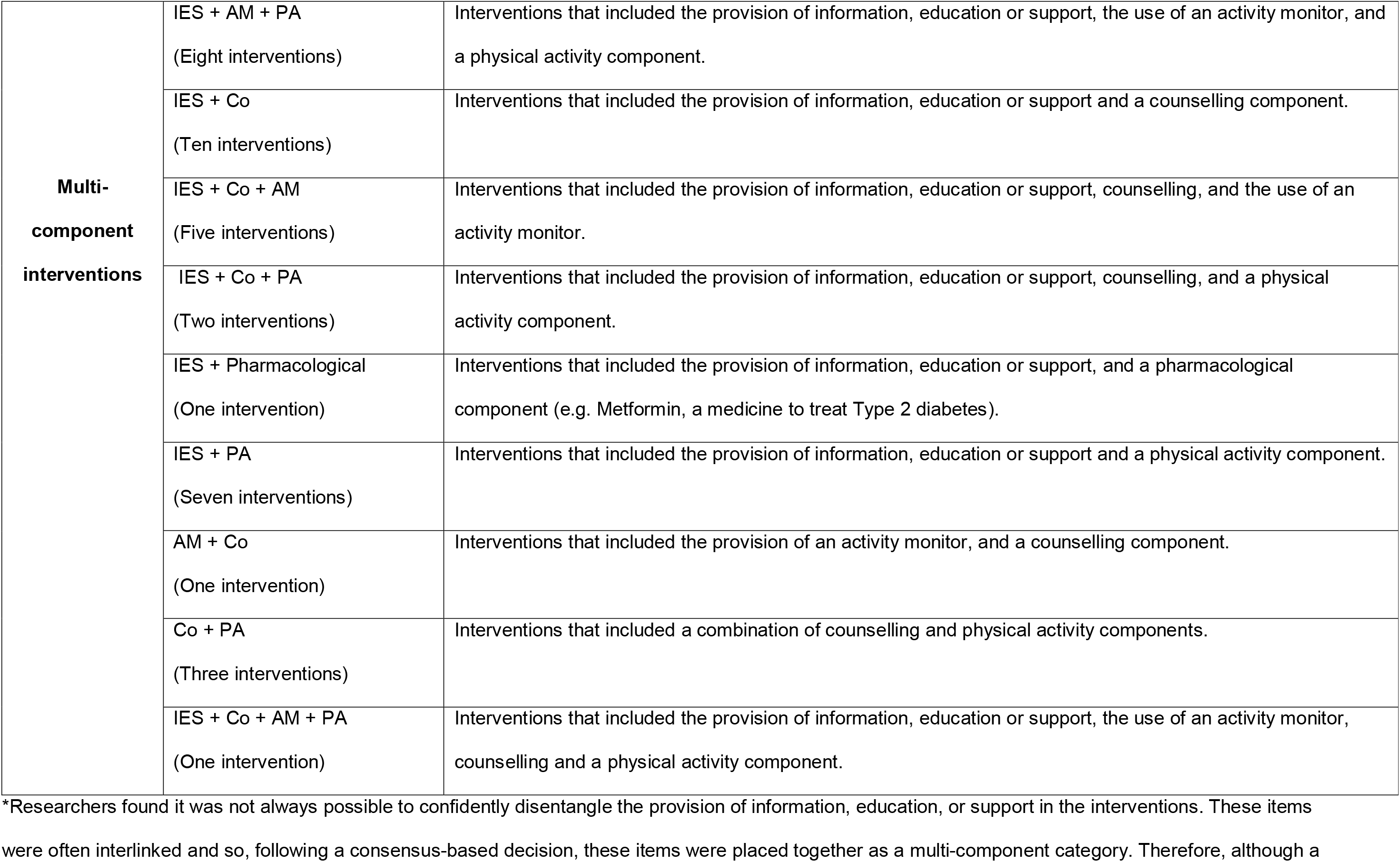
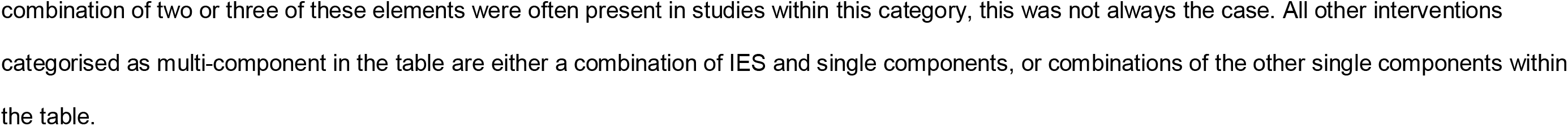
Definition of intervention categories.

### Risk of bias

Figure 2 shows a summary of the risk of bias assessment. The following domains were graded as having the largest proportion of high risk of bias: performance bias (the absence of blinding of participants and personnel), detection bias (a lack of blinding of the outcome assessment), and attrition bias (incomplete outcome data reported). Unclear risk of bias judgements were commonly applied to both aspects of selection bias (random sequence generation and allocation concealment) and reporting bias (selective reporting). We examined funnel plots for all comparisons of intervention vs. control, from which there was no evidence of publication bias. We did not examine funnel plots for individual intervention categories as there were too few studies in each group.

**Figure 2:**
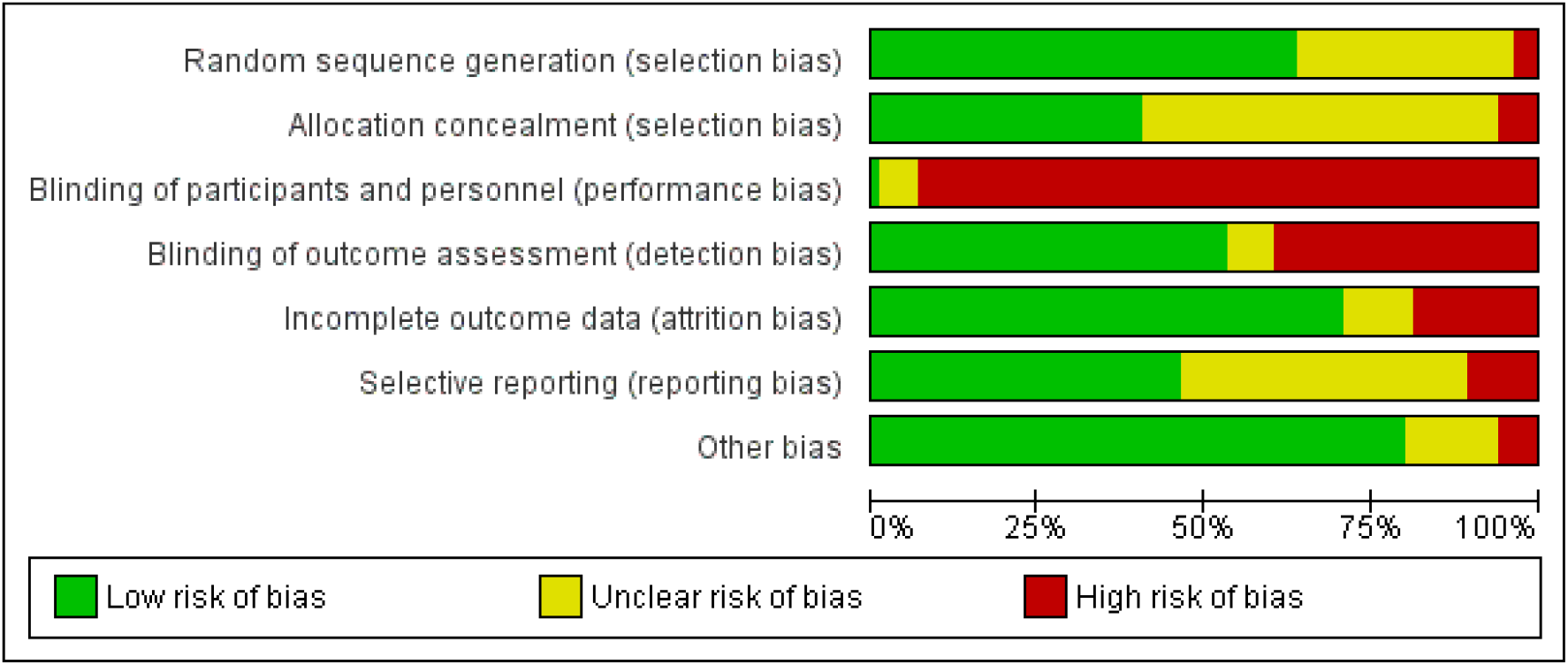
Summary of the risk of bias assessment for included studies.

### Effects of intervention categories

Studies were pooled in meta-analyses and stratified according to the outcome (time spent sedentary, or number of breaks in sedentary behaviour), how the outcome was measured (using a device, or self-report), and timing of follow-up (intervention end, or final follow-up). Analyses were conducted to explore the relative effectiveness of different intervention categories.

#### Time spent sedentary

In total, 44 studies assessed time spent sedentary using devices. Twenty-nine studies reported sufficient data to be included in the meta-analyses. Of the fourteen intervention categories, seven had results from multiple studies which were pooled, five had results from a single study and two had no relevant outcome data (see Figure 3). Only one of the pooled analyses was statistically significant; interventions that incorporated the provision of information, education, or formal support (IES), in addition to a form of counselling (Co) reduced time spent sedentary on average by MD of -52.24 minutes per day (95% CI -85.37 to -19.10, p= 0.002; I^2^= 0%; n= 170); the GRADE certainty of confidence is moderate. Two other intervention categories with results from single studies showed significant reductions in time spent sedentary: IES + Co + structured/prescribed physical activity (PA) (MD of -60 minutes per day, 95% CI -94.81 to -25.19, p= 0.0007; n= 126); the GRADE certainty of confidence is low, and Co + PA (MD of -42 minutes per day, 95% CI -59.65 to -24.35, p<0.00001; n= 300); the GRADE certainty of confidence is moderate.

**Figure 3:**
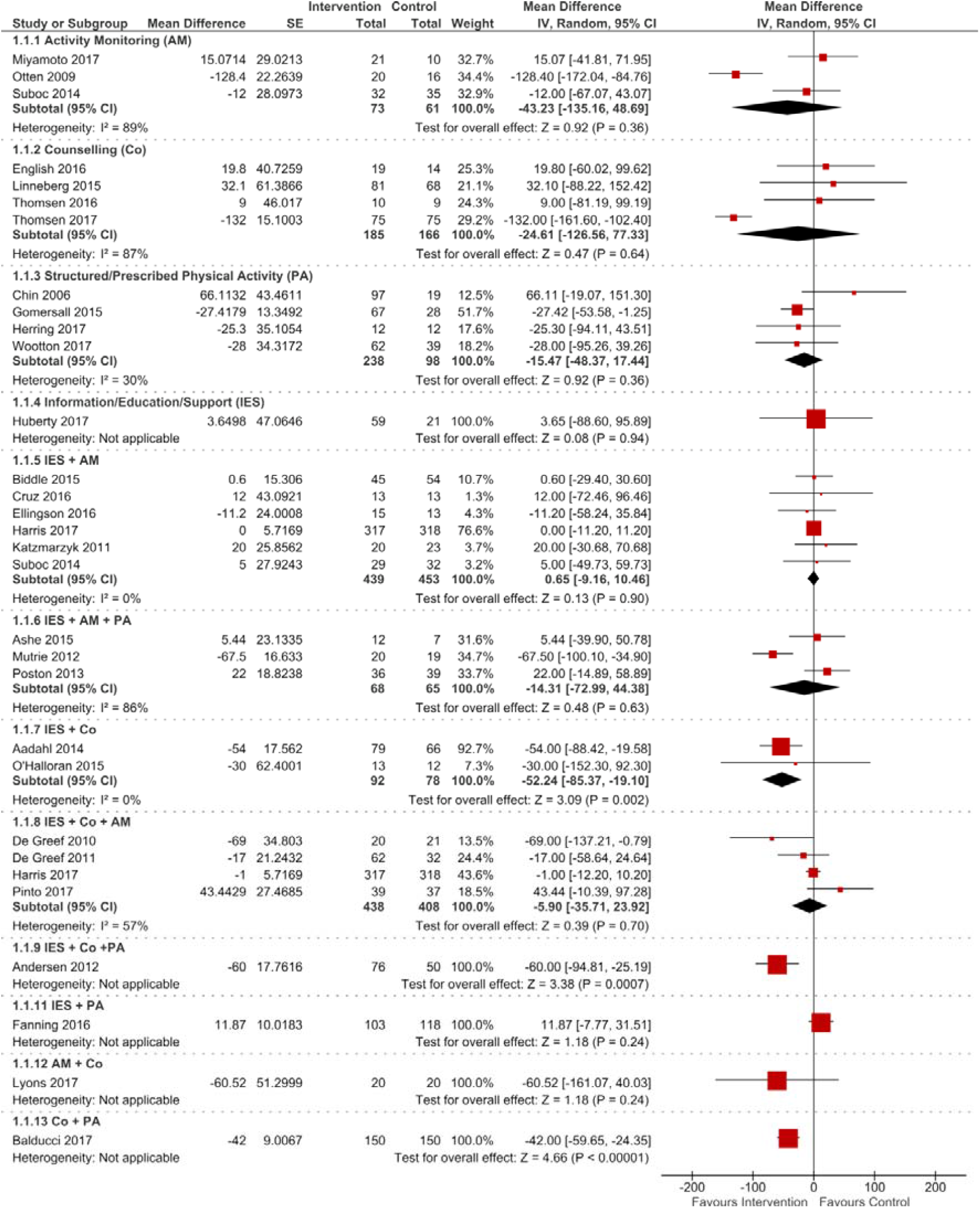
Forest plots showing effects of intervention categories to reduce time spent sedentary measured using devices (at intervention end) *. *Results for intervention categories with a minimum of two studies (7/14) were pooled. Two intervention categories (2/14) had no relevant outcome data (IES + Pharmacological; IES + Co + AM + PA).

Only 9 studies reported further follow-up beyond the end of intervention, with follow-up periods ranging from 12 to 40 weeks post-intervention. The pooled analyses showed no indication of sustained effectiveness after the end of interventions; however the single study of IES + Co + PA did show a significant reduction in time spent sedentary at 6 months post-intervention (MD of -96 minutes per day, 95% CI -131.22 to -60.78, p<0.00001; n= 97); the GRADE certainty of confidence is very low (see Figure 4).

**Figure 4:**
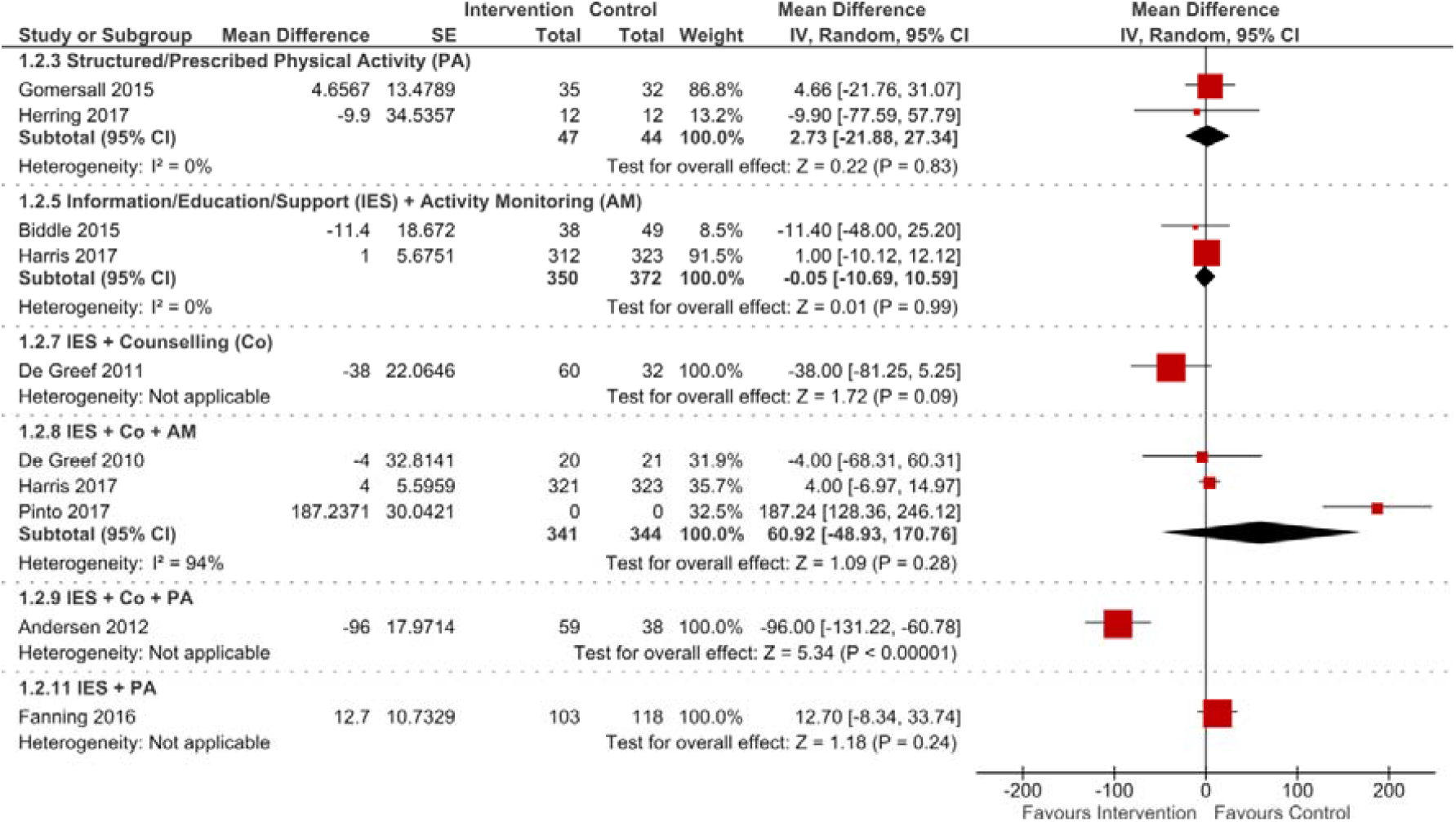
Forest plots showing effects of intervention categories to reduce time spent sedentary measured using devices (final follow-up)* *Results for intervention categories with a minimum of two studies (3/14) were pooled. Eight intervention categories (8/14) had no relevant outcome data: AM; Co; IES; IES + AM + PA; IES + Pharmacological; AM + Co; Co + PA; IES + Co + AM + PA.

In total, 40 studies assessed time spent sedentary using self-report measures. Twenty-six studies reported sufficient data to be included in the meta-analyses. Of the fourteen intervention categories, five had results from multiple studies which were pooled, four had results from a single study and five had no relevant outcome data (see Figure 5). Only one of the pooled analyses was statistically significant; interventions that incorporated the provision of information, education, or formal support, in addition to a structured/prescribed physical activity component reduced time spent sedentary on average by a significant, albeit small, effect size of SMD of -0.15 (95% CI -0.23 to -0.07; Z = 3.76, p= 0.0002; I^2^= 0%; n= 2419); the GRADE certainty of confidence is moderate.

**Figure 5:**
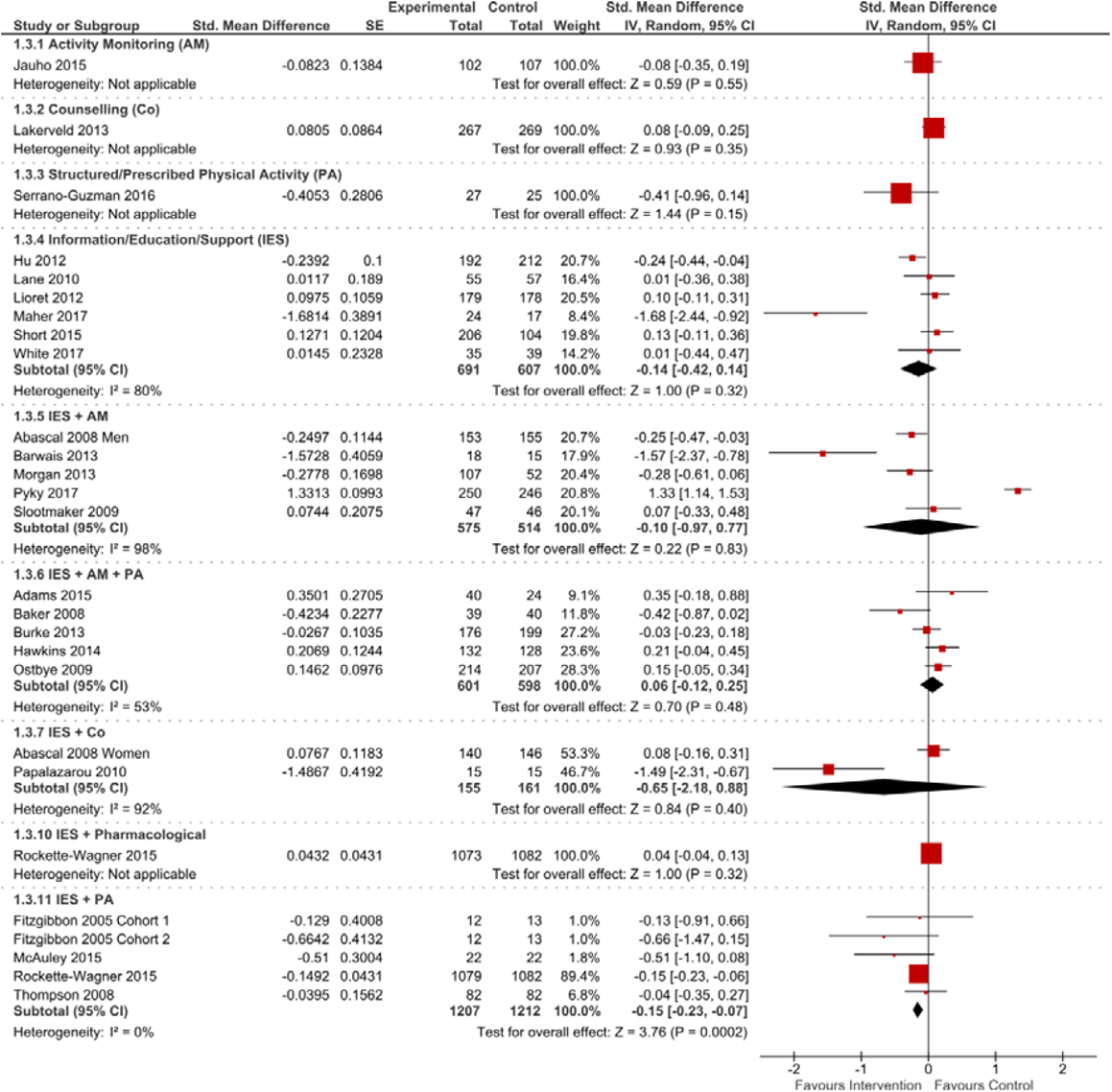
Forest plots showing effects of intervention categories to reduce time spent sedentary using self-report outcome measures (at intervention end) *. *Results for intervention categories with a minimum of two studies (5/14) were pooled. Five intervention categories had no relevant outcome data: IES + Co + AM; IES + Co + PA; AM + Co; Co + PA; IES + Co + AM + PA.

Only 5 studies reported further follow-up beyond the end of intervention, with follow-up periods ranging from 4 weeks to 18 months post-intervention. The pooled analysis showed no indication of sustained effectiveness after the end of intervention (see Figure 6); however the single study of an intervention that incorporated the provision of information, education or formal support, in addition to activity monitoring did show a statistically significant reduction in time spent sedentary at 3 months post-intervention (SMD of -0.54, 95% CI -0.88 to -0.21, p=0.002; n= 159); the GRADE certainty of confidence is low.

**Figure 6:**
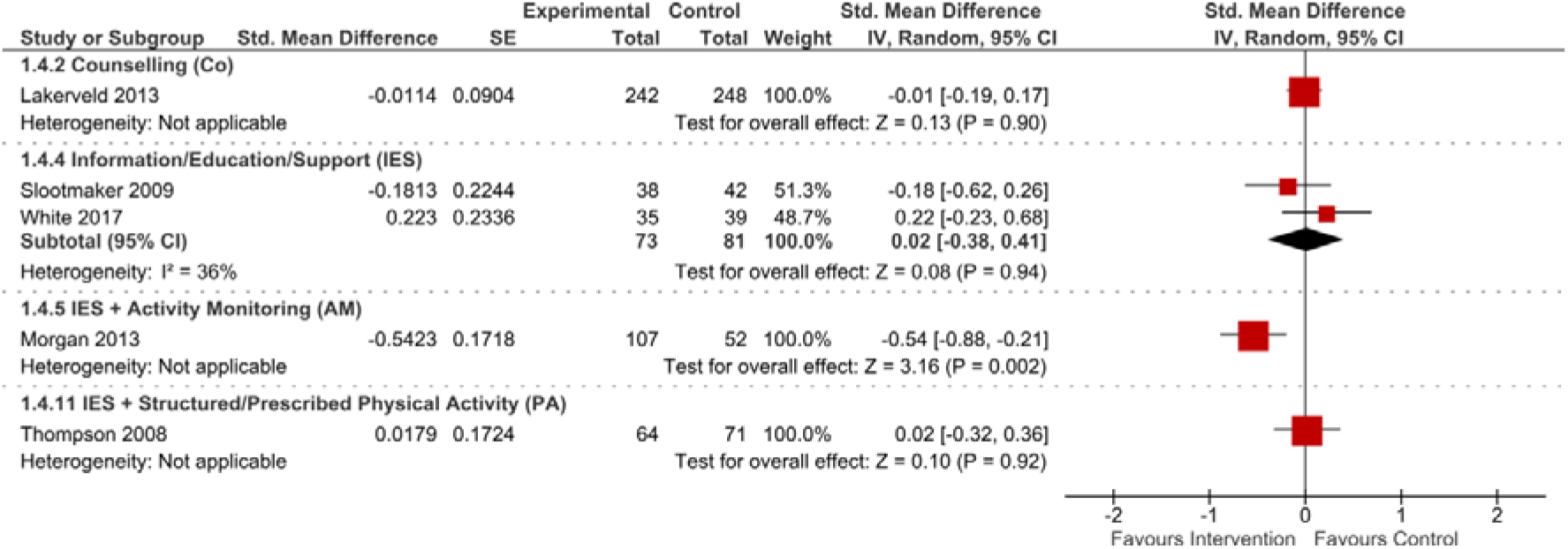
Forest plots showing effects of intervention categories to reduce time spent sedentary using self-report outcome measures (final follow-up)* *Results for intervention categories with a minimum of two studies (1/14) were pooled. Ten intervention categories had no relevant outcome data: Activity Monitoring (AM); Structure/Prescribed Physical Activity (PA); IES + AM + PA; IES + Co; IES + Co + AM; IES + Co + PA; IES + Pharmacological; AM + Co; Co + PA; IES + Co + AM + PA.

#### Number of breaks in sitting time

In total, four studies assessed the number of breaks in prolonged sitting using devices (see Figure 7). Of the fourteen intervention categories, only one (counselling) had results from multiple studies which were pooled, two had results from a single study and the remaining eleven had no relevant outcome data (see Figure 7). Counselling interventions did not show an effect in favour of the interventions (SMD= 2.18; 95% CI -1.44 to 5.80, p= 0.24; I^2^= 0%; n= 169); the GRADE certainty of confidence is moderate.

**Figure 7:**
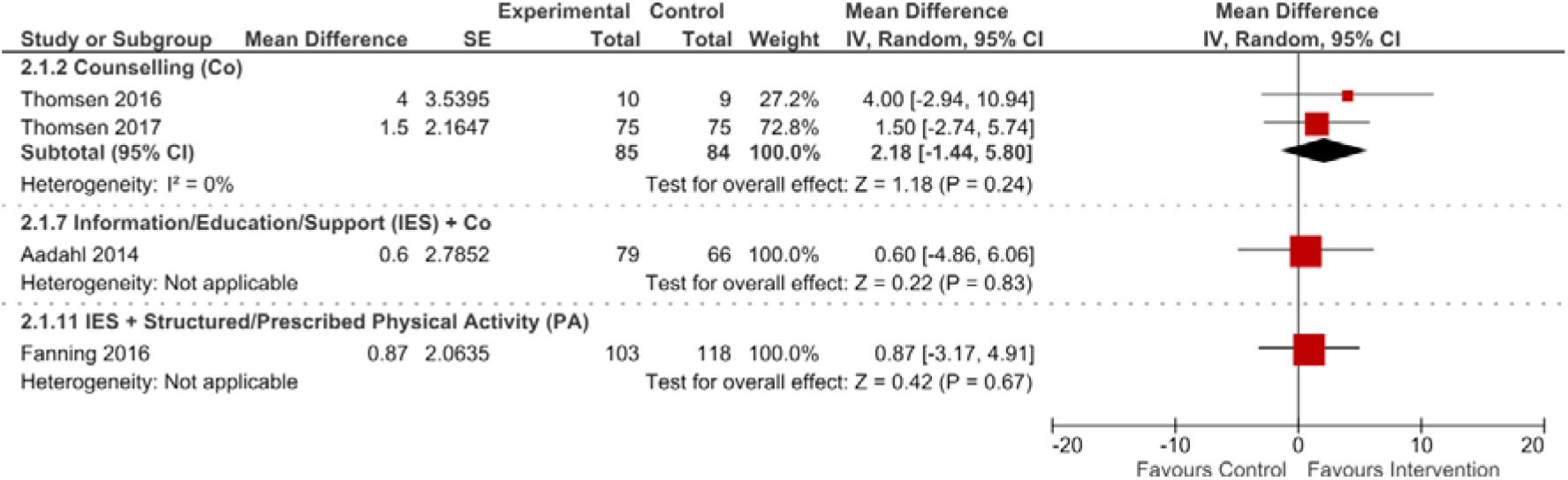
Forest plots showing effects of interventions to increase the number of breaks in sitting time using devices (at intervention end) *. *Results of intervention categories with a minimum of two studies (1/14) were pooled. Eleven intervention categories had no relevant outcome data: Activity Monitoring (AM); Structured/Prescribed Physical Activity (PA); Information/Education/Support (IES); IES + AM; IES + AM + PA; IES + Co + AM; IES + Co + PA; IES + Pharmacological; AM + Co; Co + PA; IES + Co + AM + PA.

Only one study reported further follow-up beyond the end of intervention and this did not show a significant effect (intervention incorporating information, education or formal support in addition to structured/prescribed physical activity: SMD= 2.57; 95% CI -1.85 to 6.99, p= 0.25; n= 221); the GRADE certainty of confidence is moderate.

## Discussion

This systematic review and meta-analysis explored the effects of interventions to reduce time spent sedentary and increase the number of breaks in sedentary behaviour by breaking up prolonged bouts of sitting. The majority of the results in meta-analyses were not statistically significant in favour of the interventions and most evidence was not of a high quality. Meta-analyses suggest that only interventions which focused on providing participants information, education, or formal support (such as advice or recommendations), in conjunction with either counselling (such as motivational interviewing) or with structured/prescribed physical activity (such as a walking group) led to reductions in total time spent sedentary immediately post-intervention, when measured with devices only or self-report only, respectively. Interestingly, results from a single RCT showed that an intervention which provided information, education or formal support in conjunction with counselling and with structured/prescribed physical activity also led to reductions in objectively measured total time spent sedentary immediately post-intervention and this was sustained at 6 months follow-up (114). There were very few studies, with very few participants, assessing the effect of interventions on increasing the number of breaks in sedentary behaviour; only one meta-analysis could be performed, which did not show any effectiveness of counselling interventions to increase the number of breaks in sedentary behaviour.

Evidence from preceding reviews and meta-analyses indicate the potential for interventions to reduce sedentary behaviour across a range of populations (23-27). Research suggests that reallocating 30 minutes per day of sedentary behaviour to light physical activity is clinically beneficial, with the impact even stronger when reallocating the same amount of sedentary behaviour to MVPA (127-130).

The present review is consistent with previous reviews, demonstrating that it is possible to reduce sedentary behaviour by over 30 minutes per day. In our pooled analyses we found only one intervention category reduced sedentary behaviour immediately post-intervention by more than 30 minutes per day (interventions incorporating information, education, or formal support, with counselling reduced sedentary behaviour by 52 minutes per day; measured objectively by devices). A single study reported an even larger effect for an intervention incorporating information, education, or formal support in conjunction with counselling and structured/prescribed physical activity (showing a reduction in sedentary behaviour by 60 and 92 minutes per day immediately and 6 months post-intervention, respectively) (114). We also found interventions incorporating information, education, or formal support, with a structured/prescribed physical activity component showed on average a small but statistically significant reduction in sedentary behaviour (measured by self-report measures). This finding is supported by recent work reviewing sedentary behaviour interventions in overweight and obese participants. Zabatiero et al. (131) found that education alone was not sufficient to reduce sedentary behaviour, and that the inclusion of a physical activity component was important for the success of interventions.

To our knowledge, this is the first review to utilise a rigorous process to precisely categorise interventions, in order to unpick the combination of components that could be effective for reducing sedentary behaviour. Previous meta-analyses have combined interventions according to similar outcome measures (23, 24, 28); or they have used broad categories. Peachey et al. (30) broadly categorised interventions as behavioural, environmental, or multi-component in their review; in the subgroup analyses by Shrestha et al. (28), broad categories of interventions such as educational, multi-component, TV restricting, and workplace were used. The recently published review by Blackburn et al. (29) applied similar broad categories and also examined the level of complexity of interventions, based on a number of different dimensions including number of components, targeted behaviours and level of skill required. They found that environmental interventions targeting reductions in sedentary behaviour were particularly successful. They concluded that interventions may need to be more complex to address the multi-faceted nature of sedentary behaviour, although reported that a higher level of complexity was not necessarily associated with better outcomes in terms of sustained change.

Due to the nature of the categorising of interventions, the present review had fewer studies in each meta-analysis than previous reviews with broader categories. However, the aims of the present review were to better understand the key components within the interventions, and to examine whether there were particular categories of interventions that showed effectiveness. Although two intervention categories showed benefits, the small number of studies and the quality of data limit the interpretation of findings. The categories of interventions developed during the conduct of this review provide a novel method of interpreting the sedentary behaviour literature. We hope to provide a springboard for future evidence syntheses to develop, refine and build consensus for classifying interventions which is different from categorising interventions according to behaviour change techniques.

The findings of the present review are informative for researchers, clinicians and public health professionals, but it is important to be mindful of the context of interest when translating these results into practice. When applying the evidence about the categories of interventions that showed potential, more work is required to establish the finer details of the interventions, e.g. context, mechanisms of action, and delivery for a given population. Future interventions must account for contextual factors such as the physical environment (132, 133), the social and cultural environment (134), the age of participants (18), and the types of sedentary behaviours performed and which of these sedentary behaviours would be most acceptable to change for individuals (135). Thus, future research in this area should consider the reporting of qualitative intervention development work, an improved reporting of interventions, e.g. using the Template for Intervention Description and Replication (TIDieR) checklist (136), use of outcome measures that capture the amount and types of sedentary behaviour (137, 138), and conducting process evaluations to help to disentangle why and in which contexts interventions are effective or ineffective (139).

## Strengths and limitations

Robust methods were used throughout the conduct of the review. A comprehensive search strategy was developed with input from an information specialist; two review authors independently screened search results and assessed the quality of included studies using the GRADE approach. When data were available, the estimates of the intervention effects were synthesised using meta-analyses and adjusted for clustering where necessary. However, caution should be used when interpreting the findings due to the limitations of the review, which largely relate to the nature of the evidence base. Although efforts were made to combine studies with similar outcomes, there was variation in the tools used to measure outcomes across studies, and considerable variation in the self-report outcome measures used to measure sedentary behaviour. We acknowledge that in future work, it is necessary to separately assess outcome measures, such as TV viewing time, screen time, time spent sitting, and time spent sedentary, to allow for a more accurate interpretation of findings (138). This may in part account for the heterogeneity in the present review. However, it could also be due to other forms of heterogeneity such as differences in population and intervention. The meta-analyses were limited by having a small number of studies with a small number of participants for each outcome, which precluded an examination of heterogeneity by subgroup analyses. Finally, the risk of bias assessment within studies and the GRADE assessment across outcomes showed that most of the quality of evidence was low to moderate, primarily due to relatively small sample sizes and/or wide confidence intervals indicative of imprecision of results. In addition, high risk of performance and detection bias was commonly found in the studies, because blinding was difficult in the majority of these behavioural change interventions.

## Conclusions

To our knowledge, this is the first systematic review of effects of interventions on sedentary behaviour to precisely categorise interventions according to the types of components they comprised. From the limited pool of studies, findings suggest that the most promising interventions were those that were composed of the provision of information, education, or formal support with behaviour change counselling, or the provision of information, education, or formal support and structured/prescribed physical activity. There were limited studies measuring sustained behaviour change. Future interventions would benefit from focusing on the long-term maintenance of reduced sedentary behaviour.

## Supporting information

Additional File 1: PRISMA checklist

Additional File 2: MEDLINE Search Strategy

Additional File 3: Study Characteristics Table

Additional File 4: GRADE assessment

## Data Availability

Data sharing is not applicable to this article as no datasets were generated; all data analysed were obtained from the published articles included in the review.

## Acknowledgements

We acknowledge the help and support of our Information Scientist, Deirdre Andre (DA), University of Leeds. We also acknowledge the support of Eleanor Grant (EG) and Andre Etchebarne (AE) in categorising the interventions and data extraction. We also thank Dr Mark Perry for his contributions to the design of this review.

This report presents independent research funded by the National Institute for Health Research (NIHR) under its Programme Grants for Applied Research Programme, (Development and evaluation of strategies to reduce sedentary behaviour in patients after stroke and improve outcomes, RP-PG-0615-20019).

The views expressed in this publication are those of the author(s) and not necessarily those of the NIHR or the Department of Health and Social Care.

## Supporting Information

**Additional file 1:** PRISMA checklist (DOC)

**Additional file 2:** MEDLINE search strategy (DOC)

**Additional file 3:** Table of characteristics for included studies (DOC)

**Additional file 4:** GRADE assessment (DOC)

## Notes

### Competing Interest Statement

The authors have declared no competing interest.

### Clinical Protocols

https://www.crd.york.ac.uk/prospero/display_record.php?RecordID=83751

### Author Declarations

Ethical approval was not required because this review retrieved and synthesised data from already published studies.

