## Additional File 2: MEDLINE Search Strategy for "A systematic review and meta-analysis of non-workplace interventions to reduce time spent sedentary in adults"

1	Sedentary Lifestyle/ (6891)

2	(sedentary or sedentariness or sitting or sedentarism).ti. (6158)

3	((sedentary or sitting or seated) adj5 (behavio* or lifestyle or life-style)).tw. (6787)

4	(sedentary adj3 (adult? or men or women or males or females or individuals or people or population?)).tw. (4991)

5	((inactiv* or no exercise or nonexercise or non exercise) adj3 (adult? or men or women or males or females or individuals or people)).tw. (2490)

6	((sitting or sit or seated) adj3 (task* or time or bout* or break*)).tw. (1696)

7	low energy expenditure.tw. (139)

8	(leisure time adj5 (physical* activ* or passive or inactiv*)).tw. (3352)

9	((sitting or lying) adj2 posture*).tw. (971)

10	(prolong* adj2 (reclin* or sit or sitting or seated)).tw. (511)

11	chair rise?.tw. (332)

12	((light or low) adj "physical activ*").tw. (1738)

13	(time adj3 (computer* or television or tv or video game? or videogame? or gaming or screen or media)).tw. (5495)

14	((watch* or view*) adj3 (television or tv)).tw. (3975)

15	(play* adj3 (video game? or videogame? or computer game?)).tw. (1181)

16	((computer* or television or tv or video game? or videogame? or gaming) and (sedentary or physical* activity* or sitting or seated or underactiv* or under activ*)).ti. (341)

17	or/1-16 [sedentary behaviour] (33677)

18	randomized controlled trial.pt. (497104)

19	controlled clinical trial.pt. (99247)

20	randomized.ab. (383974)

21	placebo.ab. (186868)

22	drug therapy.fs. (2116216)

23	randomly.ab. (260670)

24	trial.ab. (403701)

25	groups.ab. (1624153)

26	or/18-25 [RCT filter] (4071212)

27	17 and 26 [sedentary behaviour and rcts] (10021)

28	exp animals/ not humans.sh. (4680104)

29	27 not 28 [human only studies] (9668)

30	(exp Child/ or Adolescent/ or exp Infant/) not exp Adult/ (1827135)

31	29 not 30 [adult only studies] (8105)
