## Additional File 3: Study Characteristics Table for "A systematic review and meta-analysis of non-workplace interventions to reduce time spent sedentary in adults"

| **Authors,**  **date, Country** | **Sufficient data to be included in meta-analyses** | **Participant characteristics**  **Age (mean, (SD))**  **Gender (% female)**  **Ethnicity** | **Intervention group description**  **(Intervention category and brief intervention description)** | **Control group description** | **Outcome measures** |
| --- | --- | --- | --- | --- | --- |
| **Activity Monitoring (AM) Interventions using devices as outcome measures:**  Interventions that included a device to measure activity (including pedometers, activity monitors such as Fitbits or smartphone applications) as a key/central component and incorporated feedback, goal setting and self-monitoring due to the nature of some of the devices. | | | | | |
| Miyamoto 2017  Japan | Yes | **Age:**  N-LPA intervention: 60 (3)  LPA intervention : 62 (2)  Control: 60 (3)  **Gender:**  22%  **Ethnicity:**  Not reported | **Intervention category: AM**  **Intervention descriptions:**  **Intervention 1:**  **Non-locomotive physical activity (N-LPA)**  All patients wore accelerometers during the intervention with the display turned on so that they received visual feedback about their activity levels. N-LPA group were verbally instructed to increase their N-LPA.  **Intervention 2:**  **Locomotive physical activity group (LPA)**  All patients wore accelerometers during the intervention with the display turned on so that they received visual feedback about their activity. The L-PA group were told to increase their LPA before the start of the intervention and at the 8 and 12 week follow-up. | Participants were given no instruction regarding physical activity at the follow-up. They wore an accelerometer that measured physical activity but had the display turned off so that they did not receive visual feedback about their activity levels. | Objective  Time spent sedentary |
| Otten 2009  USA | Yes | **Age:**  Intervention: 43 (13)  Control: 42 (13)  **Gender:**  70%  **Ethnicity:**  Intervention:  95% White  Control:  94% White | **Intervention category: AM**  **Intervention description:**  A weekly limit on the participant TV time was placed. When the limit was reached, the monitor shut the TV off and would not allow it to be turned on again until the next week. | TV monitors were placed on TVs for continued observation and to control for effect of their presence. Controls were instructed to continue usual TV viewing habits. | Objective  TV viewing time |
| Suboc 2014  USA | Yes | **Age:**  Pedometer + Website: 63 (8)  Pedometer: 64 (7)  Control: 62 (7)  **Gender:**  34%  **Ethnicity:**  Not reported | **Intervention category: AM**  **Intervention description:**  Participants received a pedometer to wear with a goal to increase their physical activity by 10% each week above baseline levels. | Participants were asked not to change their behaviour during the study period. | Objective  Time spent sedentary |
| **Activity Monitoring (AM) Interventions using self-report outcome measures:**  Interventions that included a device to measure activity (including pedometers, activity monitors such as Fitbits or smartphone applications) as a key/central component and incorporated feedback, goal setting and self-monitoring due to the nature of some of the devices. | | | | | |
| Jauho 2015  Finland | Yes | **Age:**  Intervention  17.9 (0.8)  Control  18.0 (0.9)  **Gender:**  All male  **Ethnicity:**  Not reported | **Intervention category: AM**  **Intervention description:**  Participants were given a wrist-worn watch-style physical activity monitor. | Participants continued their normal life and they were given otherwise similar but blinded devices providing only the time of day. | Self-report  Time spent sitting |
| **Counselling (Co) Interventions using devices as outcome measures:**  Interventions that included counselling as part of the intervention description or a recognised type of counselling e.g. motivational interviewing, cognitive behaviour therapy.   - Not just advice; support; or giving solutions and ideas; but these components could be included as part of counselling type interventions. - This did not necessarily have to be delivered by a trained counsellor, but they would be practising recognised techniques. | | | | | |
| English 2016  Australia | Yes | **Age:**  Intervention group  65.4 (12.3)  Control  67.8 (13.8)  **Gender:**  34%  **Ethnicity:**  No reported | **Intervention category: Co**  **Intervention description:**  Four motivational interviewing sessions. | To match the groups for attention, participants received the same schedule of interviews, with a placebo message of increasing calcium for bone health. | Objective  Time spent sitting |
| Linneberg 2015 | Yes | **Age:**  Intervention: 52 (14)  Control: 52 (14)  **Gender:**  Intervention: 63%  Control: 50%  **Ethnicity:**  Not reported | **Intervention category: Co**  **Intervention description:**  Individual lifestyle counselling sessions. | Participants were instructed to maintain their usual lifestyle, including physical activity level and sedentary behaviour. | Objective  Time spent sitting |
| Thomsen 2016  Denmark | Yes | **Age:**  Intervention: 65 (9)  Control: 54 (14)  **Gender:**  60%  **Ethnicity:**  Not reported | **Intervention category: Co**  **Intervention description:**  Individual motivational counselling sessions, and individually tailored text messages. | Participants were encouraged to maintain their usual lifestyle during the intervention period. | Objective  Time spent sitting  Breaks in sitting time |
| Thomsen 2017  Denmark | Yes | **Age:**  Intervention: 60 (11)  Control: 60 (13)  **Gender:**  81%  **Ethnicity:**  Not reported | **Intervention category: Co**  **Intervention description:**  Individual motivational counselling sessions, and individually tailored text messages. | Participants were encouraged to maintain their usual lifestyle during the intervention period. | Objective  Time spent sitting  Breaks in sitting time |
| **Counselling (Co) Interventions using self-report outcome measures:**  Interventions that included counselling as part of the intervention description or a recognised type of counselling e.g. motivational interviewing, cognitive behaviour therapy.   - Not just advice; support; or giving solutions and ideas; but these components could be included as part of counselling type interventions. - This did not necessarily have to be delivered by a trained counsellor, but they would be practising recognised techniques. | | | | | |
| Lakerveld 2013  Netherlands | Yes | **Age:**  Intervention: 44 (5)  Control: 43 (6)  **Gender:**  59%  **Ethnicity:**  Not reported | **Intervention category: Co**  **Intervention description:**  Individual counselling sessions, followed by 3-monthly sessions by phone. | Participants received brochures with information and guidelines with regard to healthy physical activity levels, a healthy diet and, if relevant, smoking cessation. | Self-report  TV viewing time |
| **Structured/Prescribed Physical Activity (PA) Interventions using devices as outcome measures:**  Interventions that included structured or prescribed exercise regimes including walking, yoga, dance, gym based training programmes. These could be either group based or individual and included exercise contracts. | | | | | |
| Breyer 2010  Austria | No | **Age:**  Intervention:  61.9 (8.87)  Control:  59.0 (8.02)  **Gender:**  55%  **Ethnicity:**  Not reported | **Intervention category: PA**  **Intervention description:**  Participants trained three times/week for one hour at recommended 75% of their initial maximum heart frequency. | No intervention. | Objective  Time spent sitting |
| Chin 2006  Holland | Yes | **Age:**  Resistance training  81.0 (5.8)  Functional training  82.1 (4.9)  Combined training  80.9 (6.3)  Control  81.3 (4.4)  **Gender:**  80%  **Ethnicity:**  Not reported | **Intervention category: PA**  **Intervention descriptions:**  **Intervention 1**  **Resistance training**  Resistance training was performed twice a week during six months. Sessions lasted 45–60 minutes.  **Intervention 2**  **Functional-skills training**  Functional-skills training was performed twice a week during six months.  **Intervention 3**  **Combination**  Participants performed once weekly the resistance training and once weekly the all-round functional-skills training protocol. | The control group was designed to provide attention, social interaction. Participants were told that they were assigned to an 'educational' program (i.e. group discussions about topics of interest to older people such as history of the 20^th^ century, music). | Objective  Time spent sitting |
| de Roon 2017  Holland | No | **Age:**  Control group: 60.0 (4.9)  Diet group: 60.5 (4.6)  Mainly exercise group: 59.5 (4.9)  **Gender:**  All female  **Ethnicity:**  Not reported | **Intervention category: PA**  **Intervention description:**  An exercise training programme. The main emphasis was placed on the exercise programme, which contains four hours of MVPA/week in group- and individual sessions. | Participants were requested to keep their weight stable by adhering to the baseline diet, and maintaining their habitual exercise pattern. | Objective  Time spent sitting |
| Gomersall 2015  Australia | Yes | **Age:**  Moderate intervention  41 (12)  Extensive intervention group  45 (10)  Control  43 (10)  **Gender:**  63%  **Ethnicity:**  Not reported | **Intervention category: PA**  **Intervention descriptions:**  **Intervention 1**  **Moderate intensity**  The goal of the programme was to increase MVPA by 150 minutes/week. Half of the prescribed physical activity was to be accumulated in structured, supervised group classes, and half in the participants’ own time using modalities of their choice.  **Intervention 2**  **Extensive intensity**  The goal of the programme was to increase MVPA by 300 minutes/week in the Extensive group. Half of the prescribed physical activity was to be accumulated in structured, supervised group classes, and half in the participants’ own time using modalities of their choice. | Participants were not given any instructions about what they should do. | Objective  Time spent sedentary |
| Helgadottir 2017  Sweden | No | **Age:**  Light exercise:  43.5 (10.7)  Moderate exercise:  45.7 (12.1)  Vigorous exercise:  46.2 (13.2)  **Gender:**  66.3%  **Ethnicity**  Not reported | **Intervention category: PA**  **Intervention descriptions:**  **Intervention 1:**  **Light exercise**  This consisted of yoga classes (or similar) with a focus on gentle stretching and controlled breathing. All exercise sessions were 60 min duration and participants were requested to participate in the exercise classes three times per week.  **Intervention 2:**  **Moderate exercise**  This consisted of an intermediate-level aerobics class; and ‘vigorous exercise’, a higher intensity aerobics/strength-training and balance class. All exercise sessions were 60 min duration and participants were requested to participate in the exercise classes three times per week.  **Intervention 3:**  **Vigorous exercise**  This consisted of a higher intensity aerobics/strength-training and balance class. All exercise sessions were 60 min duration and participants were requested to participate in the exercise classes three times per week. | Participants received standard treatment for depression administered by their primary care physician who was responsible for determining the type of treatment. In many cases, this consisted of counselling with a CBT focus. | Objective  Time spent sedentary |
| Herring 2017  UK | Yes | **Age:**  Intervention  44.3 (7.9)  Control  52.4 (8.1)  **Gender:**  91.7%  **Ethnicity:**  Not reported | **Intervention category: PA**  **Intervention description:**  The intervention incorporated three 60 min gym sessions/week for 12 weeks. All sessions consisted of moderate intensity aerobic and resistance training. | Participants continued with follow up care during the 12 week exercise intervention. | Objective  Time spent sedentary |
| Wootton 2017  Australia | Yes | **Age:**  Intervention: 69 (8)  Control: 68 (9)  **Gender:**  44%  **Ethnicity:**  Not reported | **Intervention category: PA**  **Intervention description:**  Supervised, ground-based, walking training for between 30 and 45 min duration, two or three times/week. | Participants did not participate in any exercise training and were not given any instructions regarding exercise or daily physical activity. | Objective  Time spent sedentary |
| **Structured/Prescribed Physical Activity (PA) Interventions using self-report outcome measures:**  Interventions that included structured or prescribed exercise regimes including walking, yoga, dance, gym based training programmes. These could be either group based or individual and included exercise contracts. | | | | | |
| Serrano-Guzman 2016  Spain | Yes | **Age:**  Intervention: 70 (4)  Control: 70 (3)  **Gender:**  All female  **Ethnicity:**  Not reported | **Intervention category: PA**  **Intervention description:**  The intervention consisted of 24 dance sessions, three times/week. | Participants were instructed to follow the physical activity recommendations outlined in the protocols for the reduction of cardiovascular risk. During the initial examination, each woman was given a booklet stressing the importance of exercise, highlighting the risk posed by a sedentary lifestyle. | Self-report  Time spent sitting |
| **Information/Education/Support (IES) Interventions using devices as outcome measures:**  Interventions that include information, education, or support in the description or had the following features:  The intervention informed/instructed, educated or supported participants by providing advice, solutions, or recommendations. This could be in the form of something tangible e.g. a booklet, newsletter, text messages, an online app, or verbally in a one-to-one or group settings. | | | | | |
| Carlson 2012  USA | No | **Age:**  Intervention  Age range 18- 56 years  Control  Age range 18-56 years  **Gender:**  52%  **Ethnicity:**  **Intervention**  White non-Hispanic 69.9%  Hispanic 6.7%  Asian 4.9%  Black 4.9%  **Control**  White non-Hispanic 69.8%  Hispanic 7.9%  Asian 3.2%  Black 4.9% | **Intervention category: IES**  The men’s and women’s interventions were conducted separately but were very similar.  **Intervention description:**  Participants completed monthly web-based activities including learning about and applying a new behavioural skill and were encouraged to report progress on their goals and set new goals on a weekly basis. Women received monthly telephone calls from study counsellors and men received calls approximately every three months. | The control condition in the women’s study was a wait-list. In the men’s study, the control condition had access to a website that contained general health information topics (e.g., information on sun exposure protection and worksite injury prevention). | Objective  Time spent sedentary |
| Huberty 2017  USA | Yes | **Age:**  Standard:  30.83 (5.22)  Plus one:  31.05 (5.52)  Plus six:  31.48 (5.44)  Plus six choice:  31.44 (4.16)    **Gender:**  All female  **Ethnicity:**  Hispanic: 2%  Non- Hispanic: 98% | **Intervention category: IES**  **Intervention descriptions:**  Text4baby (T4b) is a free, nation-wide, mobile health information service that delivers health-related SMS to pregnant women and during the first year postpartum.  **Intervention 1**  Two physical activity SMS and one T4b SMS on Monday, Tuesday, Wednesday.  **Intervention 2**  Six physical activity SMS and one T4b SMS from Sunday to Saturday.  **Intervention 3**  Six physical activity SMS and one T4b SMS from Sunday to Saturday at a time of day the participant chooses. | Usual care (standard T4b SMS content) | Objective  Time spent sedentary |
| King 2016  USA | No | **Age:**  Affect:  59.5 10.0  Analytic:  59.5 9.5  Social:  57.9 7.7  Control:  62.8 9.8  **Gender:**  75% | **Intervention category: IES**  **Intervention descriptions:**  **Intervention 1:**  **The social mobile application**  This application included a live wallpaper display of individual avatars representing the user and other study participants. Feedback relating to how active/sedentary the user was during the day was displayed in tandem with feedback reflecting the user's group as a whole.  All personal data were displayed in reference to group averages.  **Intervention 2:**  **The affective mobile application**  This application included an avatar used to reflect how active the user was throughout the day. Positive reinforcement occurred in two ways. First, whenever a person reached pre-specified thresholds related to physical activity or trajectories of sedentary behavior.  **Intervention 3:**  **The analytic application**  This application emphasised personalised and quantitative goal-setting, behavioural feedback, informational tips promoting behaviour change, and problem-solving strategies. | Control (a dietary tracking mobile application). | Objective  Time spent sedentary |
| **Information/Education/Support (IES) Interventions using self-report measures:**  Interventions that include information, education, or support in the description or had the following features:  The intervention informed/instructed, educated or supported participants by providing advice, solutions, or recommendations. This could be in the form of something tangible e.g. a booklet, newsletter, text messages, an online app, or verbally in a one-to-one or group settings. | | | | | |
| Cotten 2016  Canada | No | **Age:**  Intervention  21.37 (3.60)  Control  21.02 (4.76)  **Gender:**  74%  **Ethnicity:**  Not reported | **Intervention category: IES**  **Intervention description:**  Text messages twice daily, one in the morning or early afternoon and one in the evening. They received one fact about sedentary behaviour at the beginning of each week and included different health risks outlined. They then received various challenges, tips, and reminders throughout the week. | The group received daily text messages in the evenings about random health or nutrition facts. | Self-report  Breaks in sitting time |
| Hansen 2012  Denmark | No | **Age:**  Intervention: 50.7 (13.6)  Control group  50.4 (13.7)  **Gender:**  65%  **Ethnicity:** not reported | **Intervention category: IES**  **Intervention description:**  The intervention website was structured as three major parts: (1) a personal page, which included individually tailored physical activity advice and a personal profile, (2) a page with training programmes and general recommendations, and (3) a forum and discussion page for questions from participants. | No intervention. | Self-report  Time spent sitting |
| Hu 2012  China | Yes | **Age:**  Intervention: 32.3 (3.5)  Control:  32.4 (3.6)  **Gender:**  All female  **Ethnicity:**  Not reported | **Intervention category: IES**  **Intervention description:**  Participants met one-on-one with a dietitian who instructed participants on how to achieve: weight reduction, dietary intake, and MVPA goals. | The group will be educated regarding general principles of healthy lifestyle that benefits  Type 2 diabetes and obesity prevention, and also informed about the current evidence showing that the lifestyle intervention is effective in women at high risk for Type 2 diabetes. | Self-report  Time spent sitting |
| Lane 2010  Ireland | Yes | **Age:**  84% of sample aged between 21 and 49 years  **Gender:**  100%  **Ethnicity:**  Not reported | **Intervention category: IES**  **Intervention description:**  Booklets were provided containing information and strategies designed to facilitate physical activity. | A ‘placebo treatment’ was delivered to the group who were mailed a healthy eating and nutrition booklet developed by the Irish Heart Foundation. | Self-report  Time spent sitting |
| Lioret 2012  Australia | Yes | **Age:**  Intervention: 33 (4)  Control: 32 (4)  **Gender:**  100%  **Ethnicity:**  Not reported | **Intervention category: IES**  **Intervention description:**  Parenting skills and behaviours that aimed to promote the development of healthy eating and physical activity behaviours in infants, along with reduced sedentary behaviours. Using group discussion, peer support, visual and written messages, and mail-outs. | Received usual care, and newsletters regarding generic issues in child. Researchers met with the participants three times over the course of the study to collect data. | Self-report  Time spent sedentary |
| Maher 2017  USA | Yes | **Age:**  Sample: 77 (9)  **Gender:**  Sample: 91%  **Ethnicity:**  Sample:  93% White  5% Asian  2% Mixed race | **Intervention category: IES**  **Intervention description:**  Video and group discussion focused on reducing sedentary behaviour. | Video and group discussion focused on social isolation as the topic for the group. This involved watching video segments and participating in group discussions. | Self-report  Time spent sedentary |
| McGuire 2001  USA | No | **Age:**  Sample: 35 (6)  **Gender:**  Sample: 79%  **Ethnicity:**  Sample:  89% White | **Intervention category: IES**  **Intervention description:**  Monthly newsletters emphasising self-weighing, increased servings of fruits and vegetables, decreased servings of high-fat foods, and walking. | This group that did not receive any treatment information. | Self-report  TV viewing time |
| Short 2015  Australia | Yes | **Age:**  Tailored intervention: 56 (Range 34-74)  Targeted intervention : 55 (Range 36-82)  Control: 55 (Range 33-75)  **Gender:**  100%  **Ethnicity:**  Not reported | **Intervention category: IES**  **Intervention descriptions:**  **Intervention 1:**  **Tailored-print intervention**  Computer-tailored newsletters. Each provided advice and feedback unique to the individual. The advice participants receive were tailored.  **Intervention 2:**  **Targeted-print intervention**  An exercise guidebook developed for promoting physical activity among breast cancer survivors. | Received the brochure ‘An active way to better health’ describing the national physical activity guidelines for Australian adults. | Self-report  Time spent sitting |
| White 2017  UK | Yes | **Age:**  Intervention: 68 (4)  Control: 69 (4)  **Gender:**  59%  **Ethnicity:**  Intervention:  96% White  2% Asian  2% Mixed or other  Control:  98% White  2% Mixed or other | **Intervention category: IES**  **Intervention description:**  A printed A5-sized information booklet outlining the health impact of sedentary behaviour and physical activity and tips on reducing sedentary behaviour and forming physical activity habits. | An factsheet was provided outlining:  The health consequences of physical activity and sedentary behaviour, and describes recommendations for duration, frequency and intensity of physical activity, and suggests that sedentary time is minimised, with examples. | Self-report  Time spent sedentary |
| **Information/Education/Support (IES) + Activity Monitoring (AM) Interventions using devices as outcome measures:**  Interventions that included the provision of information, education or support and the use of an activity monitor. | | | | | |
| Biddle 2015  UK | Yes | **Age:**  Intervention  32.4 (5.4)  Control  33.3 (5.8)  **Gender:**  68%  **Ethnicity**  20% black and minority ethnic group | **Intervention category: IES + AM**  **Intervention description:**  A 3-hour group-based structured education workshop. In addition, participants were given a sedentary behaviour and physical activity self-monitoring device. | Received an information leaflet focusing on key illness perceptions of being at risk of Type 2 diabetes, the importance of increasing physical activity and decreasing sedentary behaviour. | Objective  Time spent sedentary |
| Cruz 2016  Portugal | Yes | **Age:**  Intervention  68.8 (8.2)  Control  64.1 (8.2)  **Gender:**  13%  **Ethnicity:**  Not reported | **Intervention category: IES + AM**  **Intervention description:**  Participants received pulmonary rehabilitation and a physical activity intervention. Participants were given a pedometer and a diary to record daily steps in order to establish their baseline steps. Participants received a health contract. A physiotherapist provided feedback on performance and helped define the next short-term goal. | Pulmonary rehabilitation | Objective  Time spent sedentary |
| Ellingson 2016  USA | Yes | **Age:**  Sedentary feedback group  20.4 (1.5)  Minimal education control  19.8 (1.5)  **Gender:**  50%  **Ethnicity:**  79% white | **Intervention category: IES + AM**  **Intervention description:**  Participants received real-time feedback every other week in the form of a small vibration provided by activPAL monitors when sedentary time exceeded 30 minutes. They were given information about developing new habits to reduce sedentary time. | Minimal education regarding the risks of a sedentary lifestyle. | Objective  Time spent sedentary |
| Harris 2017  UK | Yes | **Age:**  45-54y: 33%  55-64y: 38%  65-75y: 29%  **Gender:**  36%  **Ethnicity:**  White: 80.3%  Black/African/Caribbean/Black British: 10%  Asian/Asian British: 7%  Other: 3% | **Intervention category: IES + AM**  **Intervention description:**  Key intervention components were as follows: pedometers; patient handbook; physical activity diary. | Control group participants were not provided with any feedback on their physical activity levels and did not receive materials promoting physical activity. | Objective  Time spent sedentary |
| Katzmarzyk 2011  USA | Yes | **Age:**  Control:  50.3 (7.7)  Education and pedometer:  52.7 (8.8)  **Gender:**  84%  **Ethnicity:**  72% white | **Intervention category: IES + AM**  **Intervention description:**  This group received the same educational materials as the control group, in addition to a pedometer. Participants walked outside with an interventionist for approximately 10 minutes to build self-efficacy for walking at MVPA and to observe how quickly steps accrued. | This group received minimal education: a brochure detailing the importance of physical activity for maintaining health, the physical activity guidelines, and strategies to increase physical activity levels. | Objective  Time spent sedentary |
| Suboc 2014  USA | Yes | **Age:**  Pedometer + Website: 63 (8)  Pedometer: 64 (7)  Control: 62 (7)  **Gender:**  34%  **Ethnicity:**  Not reported | **Intervention category: IES + AM**  **Intervention description:**  Participants received a pedometer and were additionally asked to log onto a secure website on at least a weekly basis At the end of each week, upon uploading pedometer information through a USB, graphical representations were provided of daily steps and how such corresponded with goals. Each week of the interactive programme was also guided by an ongoing discussion forum, and access to “ask the expert” for clarification. | Participants were asked not to change their behaviour during the study period. | Objective  Time spent sedentary |
| **Information/Education/Support (IES) + Activity Monitoring (AM) Interventions using self-report outcome measures:**  Interventions that included the provision of information, education or support and the use of an activity monitor. | | | | | |
| Abascal 2008  **Includes 2 trials:** Men in Motion  USA | Yes | **Age:**  Intervention:  44.9 (7.8)  Control:  42.8 (8.0)  **Gender:** All male  **Ethnicity:**  **Intervention:**  White non- Hispanic: 72.8%  Black non-Hispanic:  6.3%  Hispanic: 15.2%  **Control:**  White non-Hispanic**:**  69.1%  Black non-Hispanic:  4.1%  Hispanic: 21.2% | **Intervention category: IES + AM**  **Intervention description:**  The intervention was tailored to men by including short sessions, the use of a pedometer, work related language and graphics, and little extraneous information.  Participants completed monthly web-based activities which included learning about and applying new behavioural skills, and reading diet and physical activity topics | Participants were given access to an alternate website and encouraged to log on monthly. The website contained general health information of interest to men but not likely to lead to changes in diet or physical activity behaviours. | Self-report  Time spent sedentary |
| Barwais 2013 | Yes | **Age:**  Intervention  26.4 (3.0)  Control  29.0 (4.4)  **Gender:**  33%  **Ethnicity:**  Not reported | **Intervention category: IES + AM**  **Intervention description:**  Participants in the intervention group interacted with an online personal activity monitor. The device was designed to motivate a reduction in sedentary behaviour and increase physical activity. Goal-setting features were activated alongside simple graphs and charts to enhance the self-monitoring of energy expenditure. Weekly motivational emails were sent to participants when they achieved their goals. | Asked to follow their normal daily physical activities and sedentary behaviour routines. | Self-report  Time spent sedentary |
| Morgan 2013  Australia | Yes | **Age:**  SHED it- Online: 47 (11)  SHED it- Resources: 48 (11)  Control: 48 (11)  **Gender:**  All males  **Ethnicity:**  Not reported | **Intervention category: IES + AM**  **Intervention descriptions:**  **Intervention 1:**  **SHED-IT resources**  SHED-IT (Self-Help, Exercise and Diet) Resources Men were provided with a weight loss package, which included (1) a 25-minute SHED-IT Weight Loss DVD for Blokes; (2) a Weight Loss Handbook for Blokes and the Weight Loss Support Book for Blokes; and (3) a pedometer, tape measure for waist circumference, and a kilojoule counter book. The  **Intervention 2:**  **SHED-IT Online**  SHED-IT (Self-Help, Exercise and Diet using Internet Technology) Online group. In addition to receiving the materials from the SHED-IT Resources intervention, men were provided with a website and were instructed to use the online food and exercise diary. Participant was emailed seven individualised feedback sheets. | Wait-listed control | Self-report  Time spent sitting |
| Petersen 2012  Denmark | No | **Age:**  Intervention: Median 52 [41 to 62]*  Control: Median 52 [40 to 62]*  *[25^th^ percentile to 75^th^ percentile]  **Gender:**  Sample: 67%  **Ethnicity:**  Not reported | **Intervention category: IES + AM**  **Intervention description:**  Participants received a pedometer, a book with a pedometer-based goal-setting programme, a handout with a summary of the goal-setting programme, and a logbook for registration of the number of steps taken daily. The book included information of the health benefits of physical activity, encouraging participants to incorporate walking in their everyday life. | Received a leaflet from the National Board of Health in Denmark describing the benefits of physical activity and the national recommendations of a minimum of 30 min of physical activity/day. | Self-report  Time spent sedentary |
| Pyky 2017  Finland | Yes | **Age:**  Intervention: 18 (1)  Control: 18 (1)  **Gender:**  All males  **Ethnicity:**  Not reported | **Intervention category: IES + AM**  **Intervention description:**  Participants were given wrist-worn activity monitors, which displayed the accumulated daily MVPA. Participants were sent a text message reminder every three weeks to upload the physical activity data. Participants were given access to a novel mobile service (MOPOrtal). Participants received tailored feedback according to their personal physical activity through the MOPOrtal. | Sent a text message reminder every three weeks to upload the physical activity data.  Participants were given wrist-worn physical activity monitors (Polar Active), which displayed the time of day. Participants were sent a text message reminder every three weeks to upload the physical activity data.  Participants did not have access to the to a novel mobile service (MOPOrtal) service, and were not given any feedback on their physical activity level. | Self-report  Time spent sitting |
| Slootmaker 2009  Netherlands | Yes | **Age:**  Intervention: 33 (3)  Control: 32 (4)  **Gender:**  60%  **Ethnicity:**  Not reported | **Intervention category: IES + AM**  **Intervention description:**  Web-based tailored physical activity advice and a physical activity monitor. | Received information brochure with brief general physical activity recommendations. | Self-report  Time spent sedentary |
| **Information/Education/Support (IES) + Activity Monitoring (AM) + Structured/Prescribed Physical Activity (PA) Interventions using devices as outcome measures:**  Interventions that included the provision of information, education or support, the use of an activity monitor, and a physical activity component. | | | | | |
| Ashe 2015  Canada | Yes | **Age:**  Intervention:  64.8 (4.6)  Control:  63.1 (4.8)  **Gender:**  All female  **Ethnicity:** Not reported | **Intervention category: IES + AM + PA**  **Intervention description:**  The intervention had three main elements: group-based education and social support, individualised physical activity prescription (named Activity 4-1-1), and use of an activity monitor [Fitbit]. | Three education topics were similar to the intervention group (how to take public transportation, bone health and falls prevention, and personal safety), but  they did not receive information on the importance of  exercise or how to sustain an active lifestyle. Participants had no interactions with the exercise professionals nor did they receive Fitbit monitors | Objective  Time spent sedentary |
| Mutrie 2012  UK | Yes | **Age:**  Intervention: 72 (6)  Control: 70 (4)  **Gender:**  68%  **Ethnicity:**  Intervention:  63% White Scottish  32 White British  Control:  67% White Scottish  29% White British | **Intervention category: IES + AM + PA**  **Intervention description:**  Two 30-minute physical activity consultations were delivered individually to each participant by a practice nurse. The initial consultation aimed to increase walking participation. A 12-week individualised graduated walking programme in the form of a  specially designed booklet and pedometer was given to  participants. The walking group met twice weekly regardless  of the number of participants attending. The second consultation (12 weeks after the first) aimed to  maintain walking behaviour and prevent relapse. | Participants were asked to continue normal physical activity for the first 12 weeks of the study and then received the same 12-week intervention and the second consultation. | Objective  Time spent sedentary |
| Poston 2013  UK | Yes | **Age:**  Intervention: 31 (5)  Control: 30 (6)  **Gender:**  All female  **Ethnicity:**  Intervention  White: 55%  Black: 40%  Control  White: 57%  Black: 36% | **Intervention category: IES + AM + PA**  **Intervention description:**  Participants attended a one-to-  one appointment with the health trainer and were invited to weekly group sessions. Participants were encouraged to increase daily physical activity incrementally, setting goals of incremental step counts (monitored by pedometer) and maintaining the achieved physical activity level.  All attended routine antenatal care appointments and received advice regarding diet and physical activity. | Attended routine antenatal care appointments and received advice regarding diet and physical activity. | Objective  Time spent sedentary |
| **Information/Education/Support (IES) + Activity Monitoring (AM) + Structured/Prescribed Physical Activity (PA) Interventions using self-report outcome measures:**  Interventions that included the provision of information, education or support, the use of an activity monitor, and a physical activity component. | | | | | |
| Adams 2015  USA | Yes | **Age:**  Intervention:  56.73 (12.64)  Control:  61.38 (12.1)  **Gender:**  All female  **Ethnicity:**  Intervention  White: 90%  African-American: 10%  Control  White: 88%  African-American: 13% | **Intervention category: IES + AM + PA**  **Intervention description:**  Week one of the intervention consisted of a 30-minute presentation on sedentary behaviour, group brainstorming on alternatives to sedentary behaviour, and participation in an active stretching routine. Participants recorded their weekly pedometer steps and the use of the pedometer was reviewed. A total of 7 emails were sent to participants, including goal reminders, goal feedback, and examples of less sedentary behaviours. A video that showed peer models engaged in more physically activity options were sent to participants. | Wait list control | Self-report  Time spent sitting |
| Baker 2008  Scotland | Yes | **Age:**  Intervention:  47.3 (9.3)  Control:  51.2 (7.9)  **Gender:**  80%  **Ethnicity**  Not reported | **Intervention category: IES + AM + PA**  **Intervention description:**  Participants received a physical activity consultation and then followed a 12-week pedometer-based walking programme. The consultations were focused on the uptake of physical activity, in this context promoting increases in walking. | Control group were asked to maintain their normal walking levels | Self-report  Time spent sitting |
| Burke 2013  Australia | Yes | **Age:**  Intervention:  65.80 (2.95)  Control:  65.75 (3.19)  **Gender:**  48.2%  **Ethnicity:**  Not reported | **Intervention category: IES + AM + PA**  **Intervention description:**  The intervention comprised of a  booklet specially designed for seniors that provided physical activity and nutrition recommendations and  encouraged goal setting. The booklet was supported by an exercise chart, calendar, bi-monthly newsletters, resistance  band and pedometer, along with telephone and email contact by programme guides. | No intervention | Self-report  Time spent sitting |
| Hawkins 2014  USA | Yes | **Age:**  16-19y: 11%  20-24y: 39%  25-29y: 23%  >30y: 28%  **Gender:**  All female  **Ethnicity:**  53% Hispanic | **Intervention category: IES + AM + PA**  **Intervention description:**  The overall goal of the intervention  was to encourage pregnant women to achieve guidelines for physical activity during pregnancy. The participants were provided a pedometer and an activity diary. A questionnaire assessed the participants’ current stage of readiness for physical activity adoption, self-efficacy, decisional balance, use of cognitive and behavioural processes of change, and time spent in physical activity. In light of responses to the questionnaire, health educators discussed barriers and facilitators to adopting physical activity. | The health and wellness group received tips sheets and telephone booster calls on the same contact schedule as the intervention group; this controlled for contact time, while keeping the content of the two groups distinct. Specifically, after completion of the initial tailoring questionnaire, the health educator focused on general issues related to health and wellness during pregnancy instead of issues related to physical activity. | Self-report  Time spent sedentary |
| James 2015  Australia | No | **Age:** Intervention 56.2 (12.6)  Control  56.2 (12.6)  **Gender:**  77%  **Ethnicity:**  Not reported | **Intervention category: IES + AM + PA**  **Intervention description:**  Participants were provided with a workbook, pedometer and Gymstick™ (a lightweight graphite shaft, with elastic tubing and foot straps that provide resistance to exercise all major muscle groups). Each group-based session delivered simultaneous multiple health behaviour content covering a home-based walking programme (using a pedometer), home-based resistance training program (using a Gymstick™), and information  about healthy eating. | Wait-list control where participants attended the 8-week, 6-session intervention after completing 20-week study measures. | Self-report  Time spent sitting |
| **Information/Education/Support (IES) + Counselling (Co) Interventions using devices as outcome measures:**  Interventions that included the provision of information, education or support and a counselling component. | | | | | |
| Aadahl 2014  Denmark | Yes | **Age:**  Intervention: 52.2 (13.8)  Control: 51.8 (14.3)  **Gender:**  56%  **Ethnicity:** not reported | **Intervention category: IES + Co**  **Intervention description:**  Individual theory-based face-to-face sessions. Participants set specific individual goals for change in sedentary behaviour. Written materials containing strategies and suggestions for reduction of sitting time, were handed out to participants. | Participants were instructed to maintain their usual lifestyle from randomisation to the end of the follow-up period. | Objective  Time spent sitting |
| de Roon 2017  Holland | No | **Age:**  Control group: 60.0 (4.9)  Diet group: 60.5 (4.6)  Mainly exercise group: 59.5 (4.9)  **Gender:**  All female  **Ethnicity:**  Not reported | **Intervention category: IES + Co**  **Intervention description:**  Participants individually met a dietitian for the prescription of a calorie restricted diet. Additionally, they received interactive group sessions. The programme for these sessions was based on principles of cognitive behavioural therapy and motivational interviewing. | Participants were requested to keep their weight stable by adhering to the baseline diet, and maintaining their habitual exercise pattern. | Objective  Time spent sedentary |
| O’Halloran 2016  Australia | Yes | **Age:**  Intervention: 83 (5)  Control: 82 (6)  **Gender:**  89%  **Ethnicity:**  Not reported | **Intervention category: IES + Co**  **Intervention description:**  Received usual care in the community during the trial. This may have involved participants attending their general practitioner or a community physiotherapist when required.  In addition, the intervention group completed a telephone-based  motivational interviewing intervention. The motivational interviewing was designed to address issues associated with ambivalence about change in physical activity, low confidence  and fear of falling, which may prevent people after hip fracture from being more active. | Received usual care in the community during the trial. This may have involved participants attending their general practitioner or a community physiotherapist when required. | Objective  Time spent sedentary |
| **Information/Education/Support (IES) + Counselling (Co) Interventions using self-report outcome measures:**  Interventions that included the provision of information, education or support and a counselling component. | | | | | |
| Abascal 2008  Women in Balance  USA | Yes | **Age:**  Intervention: 40.8 (8.4)  Control:  41.6 (8.9)  **Gender:** All female  **Ethnicity:**  **Intervention:**  White non- Hispanic: 58.5%  Black non-Hispanic: 8.3%  Hispanic: 21.5%  **Control:**  White non-Hispanic**:** 63.3%  Black non-Hispanic: 6.1%  Hispanic: 19.4% | **Intervention category: IES + Co**  **Intervention description:**  The intervention included an initial web-based assessment, health behaviour counselling, follow-up intervention via the web, and telephone and email interaction with a health counsellor. Trained health counsellors sent individualised e-mails and made counselling phone calls. | The wait list control group received usual-care which consisted of previously scheduled provider visits and a standard set of materials summarising diet and activity recommendations. | Self-report  Time spent sedentary |
| Papalazarou 2010  Greece | Yes | **Age:**  Intervention: 33 (2)  Control: 33 (2)  **Gender:**  All female  **Ethnicity:**  Not reported | **Intervention category: IES + Co**  **Intervention description:**  Patients had the opportunity to discuss their thoughts on food intake and physical activity in sessions. Information was provided regarding to fat and fibre sources, nutritional value of foods and health benefits related to adopting a balanced dietary pattern and increasing physical activity. | Patients visited the Dietetics Department for assessment. During these assessment sessions general information was provided on adopting healthier eating and physical habits. | Self-report  TV viewing time |
| **Information/Education/Support (IES) + Counselling (Co) + Activity Monitoring (AM) Interventions using devices as outcome measures:**  Interventions that included the provision of information, education or support, counselling, and the use of an activity monitor. | | | | | |
| De Greef 2010  Belgium | Yes | **Age:**  Intervention  61.3 years  Control  61.3 years  **Gender:**  32%  **Ethnicity:**  Not reported | **Intervention category: IES + Co + AM**  **Intervention description:**  The intervention consisted of cognitive-behavioural group  sessions. In addition to the session, the participants received  a pedometer and a pedometer diary. They were asked to wear the pedometer and to record  their physical activity type and duration and number of steps at  the end of each day. In this way they could set their own step goals in the context of their daily routine. | Usual care | Objective  Time spent sedentary |
| De Greef 2011  Belgium | Yes | **Age:**  Average 62 (9) years  **Gender:**  31%  **Ethnicity:**  Not reported | **Intervention category: IES + Co + AM**  **Intervention description:**  The intervention was based on the  principles of cognitive-behavioural therapy, and Motivational  Interviewing. The session started with a motivational interview phase. The psychologist together with the participants made an individualised lifestyle plan. After this session patients started a telephone support programme given by the psychologist. A pedometer and pedometer diary were given to participants. | Usual care group. | Objective  Time spent sedentary |
| Harris 2017  UK | Yes | **Age:**  45-54y: 33%  55-64y: 38%  65-75y: 29%  **Gender:**  36%  **Ethnicity:**  White: 80.3%  Black/African/Caribbean/Black British: 10%  Asian/Asian British: 7%  Other: 3% | **Intervention category: IES + Co + AM**  **Intervention description:**  Key intervention components included: pedometers; a handbook; a physical activity diary; and individually tailored physical activity consultations were offered. | Participants were not provided with any feedback on their physical activity levels and did not receive materials promoting physical activity. | Objective  Time spent sedentary |
| Li 2017  Canada | No | **Age:**  Intervention: 52 (10)  Control: 59 (6)  **Gender:**  82%  **Ethnicity:**  Not reported | **Intervention category: IES + Co + AM**  **Intervention description:**  **Intervention**  Participants attended a session, where they received (1) a standardised group education  session about physical activity, (2) a Fitbit Flex, and (3) individual weekly activity counselling by telephone. | Wait-listed control | Objective  Time spent sedentary |
| Pinto 2017  USA | Yes | **Age:**  Intervention: 56 (9)  Control: 56 (11)  **Gender:**  All female  **Ethnicity:**  Intervention:  97% White  Control:  100% White | **Intervention category: IES + Co + AM**  **Intervention description:**  This group received the telephone-based physical activity intervention. The goal was to encourage participants to gradually increase the amount of physical activity to meet recommendations of > 30 minutes of MVPA on most days. Counselling focused on building a supportive relationship with participants, assessing motivational readiness, monitoring physical activity, identifying health concerns, and identifying and problem solving barriers to physical activity. All participants received a pedometer and a heart rate monitor. | 12 telephone calls to control for frequency of contact between the two study groups | Objective  Time spent sedentary |
| **Information/Education/Support (IES) + Counselling (Co) + Activity Monitoring (AM) Interventions using self-report outcome measures:**  Interventions that included the provision of information, education or support, counselling, and the use of an activity monitor. | | | | | |
| Albright 2014  USA (Hawaii) | No | **Age:**  Intervention: 31.6 (5.5)  Control: 32.1 (5.9)  **Gender:**  100%  **Ethnicity:**  Intervention:  Asian 34.6%  Hawaiian/other Pacific islander 32%  White 13.1%  Other Mixed 17.6%  Other (Black, Native American) 2.6%  Control:  Asian 33.3%  Hawaiian/other Pacific islander 31.4%  White 17.3%  Other Mixed 15.4%  Other (Black, Native American) 2.6% | Intervention category: **IES + Co + AM**  Intervention description:  **Tailored Telephone Counselling plus Website**  Participants were referred to a condition-specific tailored website and received 17 telephone calls with a counsellor who used motivational interviewing techniques to problem-solve barriers and set future physical activity goals (incrementally building up to 150 min/week of MVPA). They were issued a pedometer to track and set goals using steps (with goal of 10,000 steps a day). Tailored mom-centric PA “resource directories” and newsletters were key components of the website. | **Standard Website Only:**  Participants were referred to a condition-specific website that included links to “standard” (i.e., for males/females of any age) PA credible websites and resources on how to increase PA. | Self-report  Time spent sitting |
| **Information/Education/Support (IES) + Counselling (Co) + Structured/Prescribed Physical Activity (PA) Interventions using devices as outcome measures:**  Interventions that included the provision of information, education or support, counselling, and a physical activity component. | | | | | |
| Andersen 2012  Norway | Yes | **Age:**  Intervention:  35.7 (6.1)  Control:  39.7 (9.2)  **Gender:**  All male  **Ethnicity:**  Pakistani immigrant men | **Intervention category: IES + Co + PA**  **Intervention description:**  The intervention included structured group exercise  sessions, two group lectures, one individual counselling session,  written material and a phone call. | Offered organised exercise, one group lecture and written material after the end of the intervention. | Objective  Time spent sedentary |
| **Information/Education/Support (IES) + Counselling (Co) + Structured/Prescribed Physical Activity (PA) Interventions using self-report outcome measures:**  Interventions that included the provision of information, education or support, counselling, and a physical activity component. | | | | | |
| Ostbye 2009  USA | Yes | **Age:**  Intervention: 31 (6)  Control: 31 (5)  **Gender:**  100%  **Ethnicity:**  Intervention:  52.4% White  44.9% Black  2.7% other  Control:  52.9% White  45.3% Black  4% other | **Intervention category: IES + Co + PA**  **Intervention description:**  Healthy eating sessions and physical-activity group sessions. They were also provided with a study notebook with exercises, recipes, and other intervention-related information; and a pedometer. A sport stroller was provided to encourage walking for exercise outside of class and after the end of the intervention. | Received biweekly newsletters with general tips for postpartum mothers. | Self-report  TV viewing time |
| **Information/Education/Support (IES) + Pharmacological Interventions using self-report outcome measures:**  Interventions that included the provision of information, education or support, and a pharmacological component (e.g. Metformin, a medicine to treat Type 2 diabetes). | | | | | |
| Rockette-Wagner 2015  USA | Yes | **Age:**  Intensive condition : 51 (11)  Standard condition + metformin : 51 (10)  Control: 50 [10]  **Gender:**  68%  **Ethnicity:**  Intervention 1:  White: 53.8%  African American: 18.9%  Hispanic: 16.5%  American Indian: 5.6%  Asian: 5.3%  Intervention 2:  White: 56.1%  African American: 20.6%  Hispanic: 15.1%  American Indian: 4.8%  Asian: 3.4%  Control:  White: 54.2%  African American: 20.2%  Hispanic: 15.5%  American Indian: 5.5%  Asian: 4.5% | **Intervention category: IES + Pharmacological**  **Intervention description:**  Group courses are also offered during maintenance, with each course focusing on topics related to exercise, weight loss, or behavioural issues.  Metformin is started at a dose of 850 mg once daily and increased to 850 mg twice daily. | Participants received written information and an individual session addressing the importance of a healthy lifestyle for the prevention of Type 2 diabetes. Specifically, participants were encouraged to follow the Food Pyramid guidelines; to lose 5–10% of their initial weight; to increase their activity gradually with a goal of at least 30 min of an activity such as walking 5 days/week; and to avoid excessive alcohol intake. | Self-report  TV viewing time |
| **Information/Education/Support (IES) + Structured/Prescribed Physical Activity (PA) Interventions using devices as outcome measures:**  Interventions that included the provision of information, education or support and a physical activity component. | | | | | |
| Fanning 2016  USA | Yes | **Age:**  Intervention group  70.62 (0.40)  Control group  71.43 (0.43)  **Gender:**  77%  **Ethnicity:** not reported | **Intervention category: IES + PA**  **Intervention description:**  A DVD-delivered exercise program targeting flexibility, toning, and balance. Each session was designed to be used every other day for one month, and each one increased in complexity and difficulty relative  to previous sessions. In addition to the DVDs, participants received a yoga mat and two exercise bands. Participants completed exercise logs and data from the logs were used to provide feedback. | Wait list control | Objective  Time spent sedentary |
| **Information/Education/Support (IES) + Structured/Prescribed Physical Activity (PA) Interventions using self-report outcome measures:**  Interventions that included the provision of information, education or support and a physical activity component. | | | | | |
| Fitzgibbon 2005  Cohort 1  USA | Yes | **Age:**  Reported across both conditions for each cohort.  Cohort 1  44.4 (7.9)  Cohort 2  45.1 (6.9)  **Gender:**  All female  **Ethnicity:**  All black women | **Intervention category: IES + PA**  **Intervention description:**  The first 90-min weekly meeting was divided into a 45-min interactive didactic component and a 45-min exercise component (structured aerobics and walking). The second weekly meeting consisted of a 45-min exercise session. The primary difference in delivery between cohorts 1 and 2 was the time allocation between breast health and weight loss strategies. In cohort 1, equal time was devoted to each, and the breast health component was comprehensive. | Weekly emailed newsletters | Self-report  TV viewing time |
| Fitzgibbon 2005  Cohort 2  USA | Yes | **Age:**  Reported across both conditions for each cohort.  Cohort 1  44.4 (7.9)  Cohort 2  45.1 (6.9)  **Gender:**  All female  **Ethnicity:**  All black women | **Intervention category: IES + PA**  **Intervention description:**  The first 90-min weekly meeting was divided into a 45-min interactive didactic component and a 45-min exercise component (structured aerobics and walking). The second weekly meeting consisted of a 45-min exercise session. The primary difference in delivery between cohorts 1 and 2 was the time allocation between breast health and weight loss strategies. In cohort 2, approximately 80% of the time was spent on strategies related weight loss and 20% on breast health. | Weekly emailed newsletters | Self-report  TV viewing time |
| McAuley 2015  USA | Yes | **Age:**  Intervention: 60 (1)  Control: 60 (2)  **Gender:**  75%  **Ethnicity:**  Not reported | **Intervention category: IES + PA**  **Intervention description:**  A DVD-delivered exercise program targeting flexibility, toning, and  balance. Each session was designed to be used every other day for one  month, and each one increased in complexity and difficulty relative  to previous sessions. In addition to the DVDs, participants received a yoga mat and two exercise bands. Participants completed exercise logs and data from the logs were used to provide feedback. | Healthy aging DVD attention control | Self-report  Time spent sitting |
| McGuire 2001  USA | No | **Age:**  Sample: 35 (6)  **Gender:**  Sample: 79%  **Ethnicity:**  Sample:  89% White | **Intervention category: IES + PA**  **Intervention description:**  The group received monthly newsletters that emphasized self-weighing, increased servings of fruits and vegetables, decreased servings of high-fat foods, and walking. | No treatment control | Self-report  TV viewing time |
| Rockette-Wagner 2015  USA | Yes | **Age:**  Intensive condition : 51 (11)  Standard condition + metformin : 51 (10)  Control: 50 [10]  **Gender:**  68%  **Ethnicity:**  Intervention 1:  White: 53.8%  African American: 18.9%  Hispanic: 16.5%  American Indian: 5.6%  Asian: 5.3%  Intervention 2:  White: 56.1%  African American: 20.6%  Hispanic: 15.1%  American Indian: 4.8%  Asian: 3.4%  Control:  White: 54.2%  African American: 20.2%  Hispanic: 15.5%  American Indian: 5.5%  Asian: 4.5% | **Intervention category: IES + PA**  **Intervention description:**  The intervention was designed to maximise success by using the following interactive interventions: training in diet, exercise, and behaviour modification skills; frequent support; diet and  exercise interventions that are flexible, and acceptable in the  specific communities in which they are implemented. The focus of the exercise intervention is a gradual increase in brisk walking or other activities of similar intensity. Two supervised group exercise sessions per week are provided to help  participants achieve their exercise goal, but participants could also achieve the exercise goal on  their own. | Participants received written information and an individual session addressing the importance of a healthy lifestyle for the prevention of Type 2 diabetes. Specifically, participants were encouraged to follow the Food Pyramid guidelines; to lose 5–10% of their initial weight; to increase their activity gradually with a goal of at least 30 min of an activity such as walking 5 days/week; and to avoid excessive alcohol intake. | Self-report  TV viewing time |
| Sazlina 2015  Malaysia | No | **Age:**  Intervention 1: Median 63 [IQR 7]  Intervention 2: Median 64 [IQR 8]  Control: Median 63 [IQR 7]  **Gender:**  Intervention 1: 39%  Intervention 2: 48%  Control: 52%  **Ethnicity:**  Not reported | **Intervention category: IES + PA**  **Intervention descriptions:**  **Intervention 1: Personalised feedback about physical activity patterns**  A 12-week unsupervised walking activity. Participants performed gradual walking activity toward the goal of 30 minutes/day on 5 days/week at moderate intensity. Participants received structured personalised feedback and usual diabetes care.  **Intervention 2: Personalised feedback about physical activity patterns combined with peer support**  The participants received support from peer mentors in addition to engaging in unsupervised walking, receiving the personalised feedback and usual diabetes care. | Usual diabetes care. | Self-report  Time spent sitting |
| Thompson 2008  USA | Yes | **Age:**  Intervention: 30 (7)  Control: 29 (7)  **Gender:**  Intervention: 100%  Control: 100%  **Ethnicity:**  Not reported | **Intervention category: IES + PA**  **Intervention description:**  The intervention consisted of discussion-format group sessions.  Sessions included learning to read food labels, strategies for choosing healthier foods when eating out or snacking, taste-testing of healthy meals, and dissemination of  inexpensive recipes for at-home preparation of foods to increase vegetable and fruit intake and  decrease saturated fats. Weather permitting, the facilitator led a 15-minute outdoor walk at the beginning of each session. | Received mailings of a Native health magazine, address change postcards, clinic visit reminders, and phone calls to schedule clinic visits | Self-report  TV viewing time |
| **Activity Monitoring (AM) + Counselling (Co) Interventions using devices as outcome measures:**  Interventions that included the provision of an activity monitor, and a counselling component. | | | | | |
| Lyons 2017  USA | Yes | **Age:**  Intervention: 61 (5)  Control: 62 (6)  **Gender:**  85%  **Ethnicity:**  Intervention:  65% White  Control:  65% White | **Intervention category: AM + Co**  **Intervention description:**  Participants received a mini tablet mobile device and an activity monitor. The tablet was preloaded with the Jawbone Up app and synced for each participant. All the participants were provided with premade accounts that existed on a “team” with all other participants. Participants set goals  for physical activity and sedentary  behaviour. Weekly telephone counselling was provided.  Each counselling call included a check-in for any adverse events, re-evaluation of weekly goals, and action planning for the next week. | Wait-listed control | Objective  Time spent sitting |
| **Counselling (Co) + Structured/Prescribed Physical Activity (PA) Interventions using devices as outcome measures:**  Interventions that include a combination of counselling and structured/prescribed physical activity components. | | | | | |
| Balducci 2017  Italy | Yes | **Age:**  Intervention:  61.0 (9.7)  Control:  62.3 (10.1)  **Gender:**  58%  **Ethnicity:**  Not reported | **Intervention category: Co + PA**  **Intervention description:**  One individual theoretical exercise counselling session plus eight individual theoretical and practical counselling sessions (twice weekly exercise sessions), once yearly for 3 years. It was designed to promote a two-step behaviour change, that is, 1) decreasing sedentary time by substituting it with a wide range of light-intensity physical activities (LPAs) and/or interrupting prolonged sitting at home or work with brief bouts of LPA and 2) gradually increasing the time spent in purposeful MVPA by reallocating time from sedentary behaviour and/or LPA | Received only general physician recommendations for increasing daily physical activity and decreasing sedentary time. | Objective  Time spent sedentary |
| **Counselling (Co) + Structured/Prescribed Physical Activity (PA) Interventions using self-report outcome measures:**  Interventions that included a combination of counselling and structured/prescribed physical activity components. | | | | | |
| Kallings 2009  Sweden | No | **Age:**  Not reported. Participants referred to as an elderly population  **Gender:**  57%  **Ethnicity:**  Not reported | **Intervention category: Co + PA**  **Intervention description:**  The intervention group received in patient-centred counselling and individualised written prescription of physical activity. | Usual care, that is, a low-intensity intervention, with one page of written general information about the importance of physical activity for health. | Self-report  Time spent sitting |
| **Information/Education/Support (IES) + Counselling (Co) + Activity Monitoring (AM) + Structured/Prescribed Physical Activity (PA) interventions using devices as outcome measures:**  Interventions that included the provision of information, education or support, the use of an activity monitor, counselling and a physical activity component. | | | | | |
| Melville 2015  Scotland | No | **Age:**  Intervention: 44.9 (13.5)  Control: 47.7 (12.3)  **Gender:**  Intervention: 46.3%  Control: 41.7%  **Ethnicity:**  Not reported | **Intervention category: IES + Co + AM + PA**  **Intervention description: Walk Well**  Consisted of three face-to-face physical activity consultations incorporating behaviour change techniques, written resources for participants and carers, and an individualised, structured  walking programme. Participants were provided with a pedometer  to self-monitor daily step counts against the agreed, individualised  goals. The overall aim of the programme was for participants to gradually increase their daily walking time by 30 min (equivalent to around 3000 steps) on at least five days of the week, by week 12. | Wait list control | Objective  Time spent sedentary |

| **Authors,**  **date, Country** | **Sufficient data to be included in meta-analyses** | **Participant characteristics**  **Age (mean, (SD))**  **Gender (% female)**  **Ethnicity** | **Intervention group description**  **(Intervention category and brief intervention description)** | **Outcome measures** |
| --- | --- | --- | --- | --- |
| **Multiple interventions with no control group using devices as outcome measures** | | | | |
| Barone-Gibbs (2017)  USA | No | **Age:**  Sit less:  68.5 (6.7)  Get active:  67.3 (6.5)  **Gender:**  71%  **Ethnicity:**  165 not white but no further information provided | **Intervention 1 category:** **AM + Co**  **Intervention 1:**  The *Sit Less* group had a goal to reduce sedentary time by 1 hr each day. Participants received a combination of individual, in-person, and phone consultations with an exercise physiologist coupled with the use of the BodyMedia SWA activity monitors.  **Intervention 2 category: AM + Co**  **Intervention 2**  The *Get Active* group had a goal to reach 150 min of MVPA each week, accumulated in bouts of ≥10 min. Participants received a combination of individual, in-person, and phone consultations with an exercise physiologist coupled with the use of the BodyMedia SWA activity monitors | Objective  Time spent sedentary |
| Cooper 2014  UK | No | **Age:**  Men:  60.2 (7.4)  Women:  60.5 (7.4)  **Gender:**  37%  **Ethnicity:** Not reported | **Intervention 1 category: IES**  **Intervention 1:**  This group received the same treatment as in the intensive arm plus a facilitator-led, individually tailored behaviour change intervention. The behaviours targeted in the intervention were physical activity, dietary intake, medication adherence and smoking cessation.  **Intervention 2 category: IES**  **Intervention 2:**  1. A session for primary care teams to describe the treatment algorithms and targets, patient materials, and present evidence for intensive treatment.  2. Treatment algorithms based on trial data demonstrating the benefits of intensive treatment of several risk factors in people with diabetes.  3. Targets: HbA_1c_ <7%, blood pressure ≤135/85 mmHg, total cholesterol <5 mmol/l or < 4.5 mmol/l for people with a history of ischaemic heart disease.  4. Interactive audit and feedback sessions.  5. Provision of glucometers for patients.  6. Theory-based diabetes education for patients.  7. Funding to support frequent contact between patients and practitioners.  8. GPs referred newly diagnosed patients to dieticians. | Objective  Time spent sedentary |
| Joseph 2015 | No | **Age:**  Facebook group:  35.6 (6.2)  Print group:  35.3 (3.8)  **Gender:**  All female sample  **Ethnicity:**  All African American | **Intervention 1 category: IES + PA**  **Intervention 1:**  The Facebook and text message intervention was comprised four components designed to increase MVPA.  1) Weekly physical activity promotion materials posted on the group Facebook wall.  2) Discussion topics and participant engagement on the group Facebook wall.  3) Motivational text messages promoting physical activity.  4) Pedometer-based self-monitoring and goal-setting programme.  **Intervention 2 category: IES + PA**  **Intervention 2:**  The print-based Intervention was mailed to participants. It was comprised of four non-culturally tailored, self-help booklets promoting physical activity and health produced. The booklets provided information on risk factors for cardiovascular disease, the benefits of physical activity, tips and strategies to increase daily physical activity, and encouraged participants to perform a minimum of 150 minutes of MVPA per week. Participants were instructed to achieve a static goal of 8,000-10,000 steps each day. | Objective  Time spent sedentary |
| Overgaard 2018  Denmark | No | **Age:**  Intervention 1: 45 (12)  Intervention 2: 46 (10)  **Gender:**  68%  **Ethnicity:**  Not reported | **Intervention 1 category: PA**  **Intervention 1:**  The Sit Less group was instructed to reduce sedentary behaviour. A list of non-sedentary activities to replace sitting activities during time at home, at work, leisure or transport was presented and handed out.  **Intervention 2 category: PA**  **Intervention 2:**  The Exercise More group was instructed to increase MVPA to at least 30 minutes/day. | Objective  Time spent sedentary |
| Noites 2015  Portugal | No | **Age:**  Home based exercise programme +education : 63 (5)  Home based exercise programme + micro current therapy + education : 59 (2)  Control: 60 (7)  **Gender:**  20%  **Ethnicity:**  Not reported | **Intervention 1 category: IES + PA**  **Intervention 1**  The exercise programme consisted of 10 exercises. The programme lasted 2 months, with a weekly frequency of 3 times. Exercise progression occurred after 4 weeks. The programme was monitored by weekly remote supervision, using text messages, telephone calls or e-mail messages, and by fortnightly meetings to address any questions. Four educational one-on-one sessions comprised of oral explanation and pamphlets with information about healthy habits, namely the benefits of Mediterranean diet, smoking cessation and physical activity.  **Intervention 2 category: IES + PA**  **Intervention 2**  The exercise programme consisted of 10 exercises. The programme lasted 2 months, with a weekly frequency of 3 times. Exercise progression occurred after 4 weeks. The programme was monitored by weekly remote supervision, using text messages, telephone calls or e-mail messages, and by fortnightly meetings to address any questions.  Microcurrent therapy was provided to participants. The microcurrent program was completed 3 times/week for 2 months, before physical exercise.  Four educational one-on-one sessions comprised of oral explanation and pamphlets with information about healthy habits, namely the benefits of Mediterranean diet, smoking cessation and physical activity.  **Intervention 3 category: IES**  **Intervention 3:**  Four educational one-on-one sessions comprised of oral explanation and pamphlets with information about healthy habits, namely the benefits of Mediterranean diet, smoking cessation and physical activity. | Objective  Time spent sedentary |
| Raynor 2013  Pilot study 1  USA | No | **Age:**  Decrease TV : 52 (10)  Increase PA : 53 (8)  **Gender:**  88%  **Ethnicity:**  Intervention 1:  White: 83%  Intervention 2:  White: 83% | **Intervention 1 category: Co + PA**  **Intervention 1:**  The intervention consisted of group meetings. Sessions covered lessons on behavioural and cognitive skills. Each session began with a group discussion on progress toward intervention goals, which was followed by a lesson and the assignment of homework that would assist participants in meeting goals. Participants were instructed to consume a standard energy- and fat-restricted diet. Participants used a diary to record their intake from food items and beverages. Diaries were turned in weekly so that written feedback on food choices, dietary goals, and other eating behaviours could be provided. Participants were instructed to gradually increase physical activity to at least 40 minutes per day, 5 days per week. Participants were encouraged to do brisk walking and accumulate time spent being physically active by engaging in multiple short bouts.  **Intervention 2 category: Co**  **Intervention 2:**  The intervention consisted of group meetings. Sessions covered lessons on behavioural and cognitive skills. Each session began with a group discussion on progress toward intervention goals, which was followed by a lesson and the assignment of homework that would assist participants in meeting goals. Participants were instructed to consume a standard energy- and fat-restricted diet. Participants used a diary to record their intake from food items and beverages. Diaries were turned in weekly so that written feedback on food choices, dietary goals, and other eating behaviours could be provided. Participants were instructed to gradually reduce their TV watching time to 10 hours per week. | Objective  TV viewing time |
| Raynor 2013  Pilot study 2  USA | No | **Age:**  Increase PA + Decrease TV : 55 (7)  Increase PA : 53 (9)  **Gender:**  79%  **Ethnicity:**  Not reported | **Intervention 1 category: Co + PA**  **Intervention 1:**  Group meetings. Sessions covered lessons on behavioural and cognitive skills. Each session began with a group discussion on progress toward intervention goals, which was followed by a lesson and the assignment of homework that would assist participants in meeting goals. Participants were instructed to consume a standard energy- and fat-restricted diet. Participants used a diary to record their intake from food items and beverages. Diaries were turned in weekly so that written feedback on food choices, dietary goals, and other eating behaviours could be provided. Participants were instructed to gradually increase PA to at least 40 mins/day, 5 days/week. Participants were encouraged to do brisk walking and accumulate time spent being physically active by engaging in multiple short bouts. Additionally, participants were given a motion sensor to wear to measure number of steps and time spent engaged in MVPA. Mins of MVPA were recorded in the same diary in which consumption was recorded.  **Intervention 2 category: Co + PA**  **Intervention 2:**  Same as Intervention 1 + Participants were instructed to gradually reduce their TV watching time to 10 hours per week. Additionally, the overall amount of reduction in TV watching from baseline to meet the 10 hours per week goal was calculated, and in the first week, home visits occurred so that the code that the participants used to watch TV on the TV Allowances was set to limit TV watching by 50% of the overall reduction needed to meet the 10 hour per week goal. Participants were instructed to decrease their TV watching time the remaining 50% to meet the 10 hours per week goal by other behavioural strategies. | Objective  TV viewing time |
| Recio-Rodriguez 2016  Spain | No | **Age:**  Counselling and mobile app: 51 (12)  Counselling: 52 (12)  **Gender:**  62%  **Ethnicity:**  Not reported | **Intervention 1 category: AM + Co**  **Intervention 1:**  A research nurse provided counselling in physical activity and the Mediterranean diet, with the delivery of a leaflet on the session. Participants received training in the use of a mobile phone application designed to promote the Mediterranean diet and increase physical activity.  **Intervention 2 category: Co**  **Intervention 2:**  A research nurse provided counselling in physical activity and the Mediterranean diet, with the delivery of a leaflet on the session. | Objective  Time spent sedentary |
| **Multiple interventions with no control group using self-report outcome measures** | | | | |
| Berendsen 2015  Netherlands | No | **Age:**  Intervention: 55.9 (12.3)  Control: 53.8 (12.4)  **Gender:**  Intervention: 65.2%  Control: 64.0%  **Nationality:**  Intervention:  Dutch 87.4%  Other 12.6%  Control:  Dutch 90.9%  Other 9.1% | **Intervention 1 category: IES + PA**  **Intervention 1:**  Supervised exercise programme based on ‘BeweegKuur’  Individual and group meetings with a lifestyle advisor (LSA), dietician, and intensive support from a physiotherapist.  **Intervention 2 category: IES + PA**  **Intervention 2:**  Start-up exercise programme based on ‘BeweegKuur’ – same number of meetings with lifestyle advisor and dietician as intervention 1, fewer number of meetings with physiotherapist. | Self-report  Time spent sitting |
| De Cocker 2012  Belgium | No | **Age:**  Intervention  47.2 (11.2)  Control  43.5 (9.9)  **Gender:**  66%  **Ethnicity:**  Not reported | **Intervention 1 category: IES +AM**  **Intervention 1:**  Participants received personalised feedback on their steps/day, and were provided with tips and suggestions on how they can take more steps if needed.  **Intervention 2 category: AM**  **Intervention 2:**  Participants received a pedometer. | Self-report  Time spent sitting |
| Dunn 1999  USA | No | **Age:**  Lifestyle intervention  45.9 (6.8)  Structured exercise  46.2 (6.5)  **Gender:**  50%  **Ethnicity:** Not reported | **Intervention 1 category: IES + PA**  **Intervention 1:**  Participants in the structured exercise group received an exercise prescription. Supervised sessions were offered 5 days/week. Participants met quarterly for group activities and received a monthly activities calendar and a quarterly newsletter on the benefits of physical activity.  **Intervention 2 category: IES + PA**  **Intervention 2:**  Participants were advised to accumulate at least 30 minutes moderate intensity physical activity a day. Participants were invited to group activities including mall walking, volleyball and orienteering. Participants leaned behavioural strategies related to physical activity behaviour. Participants received a monthly activities calendar and a quarterly newsletter on the benefits of physical activity. | Self-report  Time spent sitting |
| Lopez-Fontana 2009  Spain | No | **Age:**  Low carb + High fat intervention : 34 (6)  High carb + Low fat intervention : 35 (8)  **Gender:**  100%  **Ethnicity:**  Not reported | **Intervention 1 category: IES**  **Intervention 1:**  Participants were individually prescribed a low carbohydrate and high fat diet.  **Intervention 2 category: IES**  **Intervention 2:**  Participants were individually prescribed a high carbohydrate and low fat diet. | Self-report  Time spent sitting |
| Muller 2016  Malaysia | No | **Age:**  Intervention: 64 (5)  Control: 63 (4)  **Gender:**  75%  **Ethnicity:**  Not reported | **Intervention 1 category: IES + PA**  **Intervention 1:**  Participants received an exercise booklet and 5 weekly SMS text messages over 12 weeks. The content of the SMS text messages was derived from effective behavior change techniques. This booklet contained information on the benefits of exercise, some safety instructions, and descriptions of 12 age-appropriate strengthening exercises that could be executed without any specific equipment. Brief warm-up and cool-down sections were included as well. One practical exercise session was conducted during the initial home visit to ensure correct execution. Participants were advised to exercise as often as possible each week to increase health benefits, but no other formal recommendations were provided.  **Intervention 2 category: IES + PA**  **Intervention 2:**  All procedures were the same as the intervention group, except they did not receive SMS text messages during the 12-week period. | Self-report  Time spent sitting |
| Spring 2012  USA | No | **Age:**  Increase FV + Increase MVPA : 33 (11)  Decrease Sat Fat + Decrease SB: 31 (11)  Increase FV + Decrease SB : 35 (12)  Decrease sat fat + Increase MVPA : 32 (10)  **Gender:**  77%  **Ethnicity:**  Not reported | **Intervention 1 category: IES + AM**  **Intervention 1:**  Increase fruit/vegetable + Increase MVPA:  **Intervention 2 category: IES + AM**  **Intervention 2:**  Decrease Sat Fat + Decrease SB:  **Intervention 3 category: IES + AM**  **Intervention 3:**  Increase fruit/vegetable + Decrease SB:  **Intervention 4 category: IES + AM**  **Intervention 4:**  Decrease sat fat + Increase MVPA:  Treatments provided three weeks of remote coaching supported by mobile decision support technology and financial incentives. During treatment, incentives were contingent on using the mobile device to self-monitor and attain behavioural targets; during follow-up they were contingent only on recording. | Self-report  Time spent sedentary |
| Spittaels 2007  Belgium | No | **Age:**  Tailored advice + email: 39.7 (8.9)  Tailored advice: 39.3 (8.7)  Standard advice: 40.9 (8.0)  **Gender:**  Tailored advice + email: 38.8%  Tailored advice: 32.0%  Standard advice: 27.0%  **Ethnicity:**  Not reported | **Intervention 1 category: IES**  **Intervention 1: Tailored advice + email**  Tailored ‘physical activity advice’ and an ‘action plan’ delivered via a website. After having received their tailored advice, participants were further encouraged to change their behaviour by five stage-of-change targeted e-mail tip sheets during a period of 8  weeks.  **Intervention 2 category: IES**  **Intervention 2: Tailored advice**  Same as intervention 1 except no emails received.  **Intervention 3 category: IES**  **Intervention 3: Standard advice**  Received standard physical activity advice via a website (based on information present in the tailored programme) | Self-report  Time spent sitting |
| Steeves 2012  USA | No | **Age:**  TV commercial stepping : 54 (9)  TV Walking group : 50 (10)  **Gender:**  80%  **Ethnicity:**  Not reported | **Intervention 1 category: IES + PA**  **Intervention 1:**  Participants were instructed to stand and “briskly” step in place, or “briskly” walk continuously around the room/house for the duration of each commercial break during at least 90 minutes of TV programming at least 5 days/week. Participants increased their stepping by incorporating it into 30 min of TV programming per day during week 1, 60 minutes per day during week 2, and 90 minutes per day after this.  **Intervention 2 category: IES PA**  **Intervention 2:**  Participants were instructed to walk “briskly” for at least 30 min at least 5 days/week. Participants built up to walking 30 minutes/day over the first 3 weeks; increasing duration from 10 minutes/day in week 1, to 20 minutes/day in week 2, to 30 minutes/day for the remainder of this study. Participants were instructed to walk for 30 min continuously or break their walking up into bouts of at least 10 min. | Self-report  TV viewing time |
