## Additional File 4: GRADE assessment for "A systematic review and meta-analysis of non-workplace interventions to reduce time spent sedentary in adults"

| # | Intervention Category | Intervention groups vs. Control groups [Time spent sedentary (mins/day) measured using devices] | | | | | |
|---|---|---|---|---|---|---|---|
| | | 1.1 At intervention end | | | 1.2 Final follow-up | | |
| | | No of participants (studies) | Certainty of the evidence (GRADE)* | Anticipated absolute effects<br>Risk difference with Intervention groups | No of participants (studies) | Certainty of the evidence (GRADE)* | Anticipated absolute effects<br>Risk difference with Intervention groups |
| 1 | Activity Monitoring (AM) | 134<br>(3 RCTs) | ⊕⊕◯◯[ab]<br>LOW | MD -**43.23**<br>(-135.16 to +48.69) | 0 | - | - |
| 2 | Counselling (Co) | 351<br>(4 RCTs) | ⊕⊕◯◯[ab]<br>LOW | MD -**24.61**<br>(-126.56 to +77.33) | 0 | - | - |
| 3 | Structured/Prescribed Physical Activity (PA) | 336<br>(4 RCTs) | ⊕⊕◯◯[ab]<br>LOW | MD -**15.47**<br>(-48.37 to +17.44) | 91<br>(2 RCTs) | ⊕⊕⊕◯[b]<br>MODERATE | MD +**2.73**<br>(-21.88 to +27.34) |
| 4 | Information/Education /Support (IES) | 80<br>(1 RCT) | ⊕⊕⊕◯[b]<br>MODERATE | MD +**3.65**<br>(-88.6 to +95.89) | 0 | - | - |
| 5 | IES + AM | 892<br>(6 RCTs) | ⊕⊕⊕⊕<br>HIGH | MD +**0.65**<br>(-9.16 to +10.46) | 722<br>(2 RCTs) | ⊕⊕⊕⊕<br>HIGH | MD -**0.05**<br>(-10.69 to +10.59) |
| 6 | IES + AM + PA | 133<br>(3 RCTs) | ⊕⊕◯◯[ab]<br>LOW | MD -**14.31**<br>(-72.99 to +44.38) | 0 | - | - |
| 7 | IES + Co | 170<br>(2 RCTs) | ⊕⊕⊕◯[b]<br>MODERATE | MD -**52.24**<br>(-85.37 to -19.1) | 92<br>(1 RCT) | ⊕⊕◯◯[bd]<br>LOW | MD -**38.00**<br>(-81.25 to +5.25) |
| 8 | IES + Co + AM | 846<br>(4 RCTs) | ⊕⊕⊕◯[a]<br>MODERATE | MD -**5.9**<br>(-35.71 to +23.92) | 685<br>(3 RCTs) | ⊕⊕⊕◯[a]<br>MODERATE | MD +**60.92**<br>(-48.93 to +170.76) |
| 9 | IES + Co + PA | 126<br>(1 RCT) | ⊕⊕◯◯[bc]<br>LOW | MD -**60**<br>(-94.81 to -25.19) | 97<br>(1 RCT) | ⊕◯◯◯[bce]<br>VERY LOW | MD -**96**<br>(-131.22 to -60.78) |
| 10 | IES + Pharmacological | 0 | - | - | 0 | - | - |
| 11 | IES + PA | 221<br>(1 RCT) | ⊕⊕⊕◯[d]<br>MODERATE | MD -**11.87**<br>(-7.77 to +31.51) | 221<br>(1 RCT) | ⊕⊕⊕◯[b]<br>MODERATE | MD +**12.7**<br>(-8.34 to +33.74) |
| 12 | AM + Co | 40<br>(1 RCT) | ⊕⊕◯◯[bc]<br>LOW | MD -**60.52**<br>(-161.07 to +40.03) | 0 | - | - |
| 13 | Co + PA | 300<br>(1 RCT) | ⊕⊕⊕◯[b]<br>MODERATE | MD -**42**<br>(-59.65 to -24.35) | 0 | - | - |
| 14 | IES + Co + AM + PA | 0 | - | - | 0 | - | - |
| | [a] Heterogeneity suggests inconsistency of results; [b] Relatively small sample size and/or wide confidence intervals indicative of imprecision of results; [c] Risk of selection bias; [d] Risk of detection bias; [e] Risk of attrition bias; *see page 4 for definition of grades | | | | | | |

### Additional File 4: GRADE Assessment

| # | Intervention Category | Intervention groups vs. Control groups [Time spent sedentary (mins/day) measured using self-report measures] | | | | | |
|---|---|---|---|---|---|---|---|
| | | **1.3 At intervention end** | | | **1.4 Final follow-up** | | |
| | | **No of participants (studies)** | **Certainty of the evidence (GRADE)*** | **Anticipated absolute effects** | **No of participants (studies)** | **Certainty of the evidence (GRADE)*** | **Anticipated absolute effects** |
| | | | | **Risk difference with Intervention groups** | | | **Risk difference with Intervention groups** |
| 1 | Activity Monitoring (AM) | 209 (1 RCT) | ⊕⊕○○[ab] LOW | SMD -**0.08** (-0.35 to +0.19) | 0 | - | - |
| 2 | Counselling (Co) | 536 (1 RCT) | ⊕⊕○○[bc] LOW | SMD +**0.08** (-0.09 to +0.25) | 490 (1 RCT) | ⊕⊕⊕○[c] MODERATE | SMD -**0.01** (-0.19 to +0.17) |
| 3 | Structured/Prescribed Physical Activity (PA) | 52 (1 RCT) | ⊕⊕○○[bd] LOW | SMD -**0.41** (-0.96 to +0.14) | 0 | - | - |
| 4 | Information/Education /Support (IES) | 1298 (6 RCTs) | ⊕⊕○○[de] LOW | SMD -**0.14** (-0.42 to +0.14) | 154 (2 RCTs) | ⊕○○○[bce] VERY LOW | SMD +**0.02** (-0.38 to +0.41) |
| 5 | IES + AM | 1089 (5 RCTs) | ⊕○○○[abe] VERY LOW | SMD -**0.10** (-0.97 to +0.77) | 159 (1 RCT) | ⊕⊕○○[ab] LOW | SMD -**0.54** (-0.88 to -0.21) |
| 6 | IES + AM + PA | 1199 (5 RCTs) | ⊕⊕○○[de] LOW | SMD +**0.06** (-0.12 to +0.25) | 0 | - | - |
| 7 | IES + Co | 316 (2 RCTs) | ⊕○○○[bce] VERY LOW | SMD -**0.65** (-2.18 to +0.88) | 0 | - | - |
| 8 | IES + Co + AM | 0 | - | - | 0 | - | - |
| 9 | IES + Co + PA | 0 | - | - | 0 | - | - |
| 10 | IES + Pharmacological | 2155 (1 RCT) | ⊕⊕⊕○[c] MODERATE | SMD +**0.04** (-0.04 to +0.13) | 0 | - | - |
| 11 | IES + PA | 2419 (5 RCTs) | ⊕⊕⊕○[c] MODERATE | SMD -**0.15** (-0.23 to -0.07) | 135 (1 RCT) | ⊕○○○[bcd] VERY LOW | SMD +**0.02** (-0.32 to +0.36) |
| 12 | AM + Co | 0 | - | - | 0 | - | - |
| 13 | Co + PA | 0 | - | - | 0 | - | - |
| 14 | IES + Co + AM + PA | 0 | - | - | 0 | - | - |

[a] Risk of selection bias; [b] Relatively small sample size and/or wide confidence intervals indicative of imprecision of results; [c] TV minutes watched per day used as a surrogate for sedentary behaviour; [d] Risk of detection bias; [e] Heterogeneity suggests inconsistency of results; *see page 4 for definition of grades

**Additional File 4: GRADE Assessment**

| # | Intervention Category | Intervention groups vs. Control groups [Number of breaks (events) in prolonged sitting time using devices] | | | | | |
|---|---|---|---|---|---|---|---|
| | | 2.1 At intervention end | | | 2.2 Final follow-up | | |
| | | No of participants (studies) | Certainty of the evidence (GRADE)* | Anticipated absolute effects<br>Risk difference with Intervention groups | No of participants (studies) | Certainty of the evidence (GRADE)* | Anticipated absolute effects<br>Risk difference with Intervention groups |
| 1 | Activity Monitoring (AM) | 0 | - | - | 0 | - | - |
| 2 | Counselling (Co) | 169 (2 RCTs) | ⊕⊕⊕◯[a] MODERATE | MD +**2.18** (-1.44 to +5.8) | 0 | - | - |
| 3 | Structured/Prescribed Physical Activity (PA) | 0 | - | - | 0 | - | - |
| 4 | Information/Education /Support (IES) | 0 | - | **-** | 0 | - | - |
| 5 | IES + AM | 0 | - | **-** | 0 | - | - |
| 6 | IES + AM + PA | 0 | - | - | 0 | - | - |
| 7 | IES + Co | 145 (1 RCT) | ⊕⊕⊕◯[a] MODERATE | MD +**0.6** (-4.86 to +6.06) | 0 | - | - |
| 8 | IES + Co + AM | 0 | - | - | 0 | - | - |
| 9 | IES + Co + PA | 0 | - | - | 0 | - | - |
| 10 | IES + Pharmacological | 0 | - | - | 0 | - | - |
| 11 | IES + PA | 221 (1 RCT) | ⊕⊕⊕◯[a] MODERATE | MD +**0.87** (-3.17 to +4.91) | 221 (1 RCT) | ⊕⊕⊕◯[a] MODERATE | **MD +2.57** (-1.85 to +6.99) |
| 12 | AM + Co | 0 | - | - | 0 | - | - |
| 13 | Co + PA | 0 | - | - | 0 | - | - |
| 14 | IES + Co + AM + PA | 0 | - | - | 0 | - | - |
| | [a] Relatively small sample size and/or wide confidence intervals indicative of imprecision of results; *see page 4 for definition of grades | | | | | | |

### Additional File 4: GRADE Assessment

| **GRADE Working Group grades of evidence** | |
|---|---|
| ⊕⊕⊕⊕ | **High certainty:** We are very confident that the true effect lies close to that of the estimate of the effect. |
| ⊕⊕⊕◯ | **Moderate certainty:** We are moderately confident in the effect estimate: The true effect is likely to be close to the estimate of the effect, but there is a possibility that it is substantially different. |
| ⊕⊕◯◯ | **Low certainty:** Our confidence in the effect estimate is limited: The true effect may be substantially different from the estimate of the effect. |
| ⊕◯◯◯ | **Very low certainty:** We have very little confidence in the effect estimate: The true effect is likely to be substantially different from the estimate of effect. |
